# Precision Education Tools for Pediatrics Trainees: A Mixed-Methods Multi-Site Usability Assessment

**DOI:** 10.1101/2024.07.23.24310890

**Authors:** Alexander Fidel, Mark V. Mai, Naveen Muthu, Adam C. Dziorny

## Abstract

**Background:** Exposure to patients and clinical diagnoses drives learning in graduate medical education (GME). Measuring practice data, how trainees each experience that exposure, is critical to planned learning processes including assessment of trainee needs. We previously developed and validated an automated system to accurately identify resident provider-patient interactions (rPPIs). In this follow-up study, we employ user-centered design methods to meet two objectives: 1) understand trainees’ planned learning needs; 2) design, build, and assess a usable, useful, and effective tool based on our automated rPPI system to meet these needs.

**Methods:** We collected data at two institutions new to the American Medical Association’s “Advancing Change” initiative, using a mixed-methods approach with purposive sampling. First, interviews and formative prototype testing yielded qualitative data which we analyzed with several coding cycles. These qualitative methods illuminated the work domain, broke it into learning use cases, and identified design requirements. Two theoretical models**—**the Systems Engineering Initiative for Patient Safety (SEIPS) and Master-Adaptive Learner (MAL) **—**structured coding efforts. Feature-prioritization matrix analysis then transformed qualitative analysis outputs into actionable prototype elements that were refined through formative usability methods. Lastly, qualitative data from a summative usability test validated the final prototype with measures of usefulness, usability, and intent to use. Quantitative methods measured time on task and task completion rate.

**Results:** We represent GME work domain learnings through process-map-design artifacts which provide target opportunities for intervention. Of the identified decision-making opportunities, trainee-mentor meetings stood out as optimal for delivering reliable practice-area information. We designed a “mid-point” report for the use case of such meetings, integrating features from qualitative analysis and formative prototype testing into iterations of the prototype. A final version showed five essential visualizations. Usability testing resulted in high performance in subjective and objective metrics. Compared to currently available resources, our tool scored 50% higher in terms of Perceived Usability and 60% higher on Perceived Ease of Use.

**Conclusions:** We describe the multi-site development of a tool providing visualizations of log level electronic health record data, using human-centered design methods. Delivered at an identified point in graduate medical education, the tool is ideal for fostering the development of master adaptive learners. The resulting prototype is validated with high performance on a summative usability test. Additionally, the design, development, and assessment process may be applied to other tools and topics within medical education informatics.

## BACKGROUND

Experiential learning through direct patient care is an important paradigm in graduate medical education (GME) (ACGME, 2017). Readily available measures of patient exposure are necessary to quantify gaps in trainee experience (Rhee et al., 2022). Such objective tools can provide data to guide a trainee’s individualized education plan and enhance future learning (Mylopoulos et al., 2016). The Reflective Practice and Precision Education conceptual models build on the Master Adaptive Learner (MAL) model, pointing to patient exposure data powerfully informing the planning phases of trainee education (Cutrer et al., 2016; Janssen et al., 2022; Schumacher et al., 2023). Using this data, trainees may situate their experience within their cohorts, identifying targeted opportunities for learning (Janssen et al., 2022). Although manually tracked case-logs show promise in providing such data, those methods are labor-intensive and limited in their ability to scale (Langdorf et al., 1998; Sequist et al., 2005). Several technology-based systems have been developed to identify trainees’ patient experiences automatically and accurately across rotations (Levin & Hron, 2017; Mai et al., 2020; Wang et al., 2023). However, it remains unknown how to optimally deliver this information to trainees.

GME trainees need targeted delivery of specific information and knowledge to enhance their educational experience. *Educational* decision support (EDS) deals with such information (Kotsiantis, 2012). We posit that EDS systems are as necessary as clinical decision support systems (CDS) that support clinical duties. To be useful and usable, EDS, like CDS, must deliver the right information to the right subset of users, at the right time, through the right information channels, and in the most usable format (Osheroff et al., 2012). Traditional technology-centered design approaches are insufficient, with outcomes of increased complexity for users, elevated error rates, poor intention to use, and abandoned adoption (Woods & Winograd, 1997; Boy, 2017). These challenges stem from a design process beginning with interface creation to fit work as imagined. User engagement occurs late in the process, if at all. The alternative, user-centered design (UCD) approaches, begin by assessing user goals, tasks, abilities, and cognition prior to design ideation. By first identifying contextual needs, interface creation instead fits work as done, and better results follow: simplified use, low error rates, high intention, and eventually, adoption (Nguyen et al., 2023; Sauro, 2010).

The objective of this study was to follow a UCD approach to design and develop a user interface overlaying an existing automated educational decision support system, and to measure the summative usability of the design product. Specifically, we sought to: describe the trainee learning environment and overall context for system implementation; use this knowledge to inform initial designs; apply formative and summative evaluation techniques to the tool; use mixed methods for data collection, consistent with CDS and UCD standards. We hypothesized that this approach would generate a prototype, meeting acceptable benchmarks for technological acceptance, predicting eventual adoption. Additionally, this detailed approach can be generalized to other medical education applications requiring user interaction.

## METHODS

We performed a prospective mixed-methods study within three distinct phases described below. Subjects were physicians from two large academic institutions: the Children’s Hospital of Philadelphia (CHOP) and the University of Rochester Medical Center (URMC). We conducted the study over a two-year period, from June 2021 to September 2023.

### Phase 1: Work Domain Assessment

The first phase of a human-centered design process, work domain assessment, identifies individuals’ roles and details the overall context or work system in which a prototype tool would be used (Johnson et al., 2005; Ratwani et al., 2018). We began with roles from residency and fellowship programs, preferentially selecting participants with more years of experience. We conducted semi-structured interviews based on interview guides specific to each role. We designed these guides to elicit how trainees make informed choices in both planned and unplanned practice area learning in both the long and short term. Based on prior literature and domain knowledge from subject matter experts on the research team, we organized questions into themes exploring the context of planning and decision making of targeted learning, namely what informs those processes and when.

We recorded and transcribed interviews. We began analysis with descriptive coding, identifying topics in the corpus. Within a second round of provisional coding we deductively organized first round topics into a predetermined “start list” of categories. These were derived from an established framework, the PETT “scan” of the Systems Engineering Initiative for Patient Safety (Holden & Carayon, 2021, p. 101). The PETT scan decomposes healthcare work systems into four parts that form its acronym: people, environment, tools, and tasks. The framework is known to improve user center designers’ grasp of the end users in healthcare {ref?}. We created topic codes and organized them under these four categories. The research team reviewed output from the second round of coding on an ongoing basis as interviews continued. We considered saturation reached when we had consensus that no new significant topical codes could be added to the defined categories.

We then completed a third analysis cycle to generate process maps that explain contexts, conditions, interactions, and consequences of trainee planned learning. We detailed each role with at least one map where dynamic webs showed abstracted contributions to trainee planned learning. To create the maps, we used Process and Causative coding methodologies. We first coded observable activity and conceptual action, identifying sequential actions, grouping them, and plotting them in maps. We also coded changes to, or occurrences of, topics from the PETT scan that influenced action. Where possible, mental models were coded describing participant explanations of decision causes. Mental model representations show trainees’ new or changing intentions, choices, objectives, values, perspectives, needs, desires, and agency.

### Phase 2: Formative Testing

Interviews uncovered a single key interaction of information gathering and critical decision making. Where that interaction occurs, predictive displays with visual representations of past and potential future patient exposure could be high impact planning aids. We chose these interactions as the context for tool use.

To support this key interaction, we analyzed transcripts, abstracting from the coded domain descriptions to questions our users ask themselves. The answers determine decisions in planned learning related specifically to this key interaction. We recoded the transcripts a final time using magnitude coding, assigning each question a numerical value reflecting the impact an informed answer would have to planned learning. The impact score, from 1 to 5 (with 5 reflecting high impact), combined rater assessments of utility of the described learnings and degrees of association with the key scenario. We developed this list of questions, information required to answer them, and their impact score into a feature prioritization matrix (FPM) (see supplementary materials).

**Figure 1:**
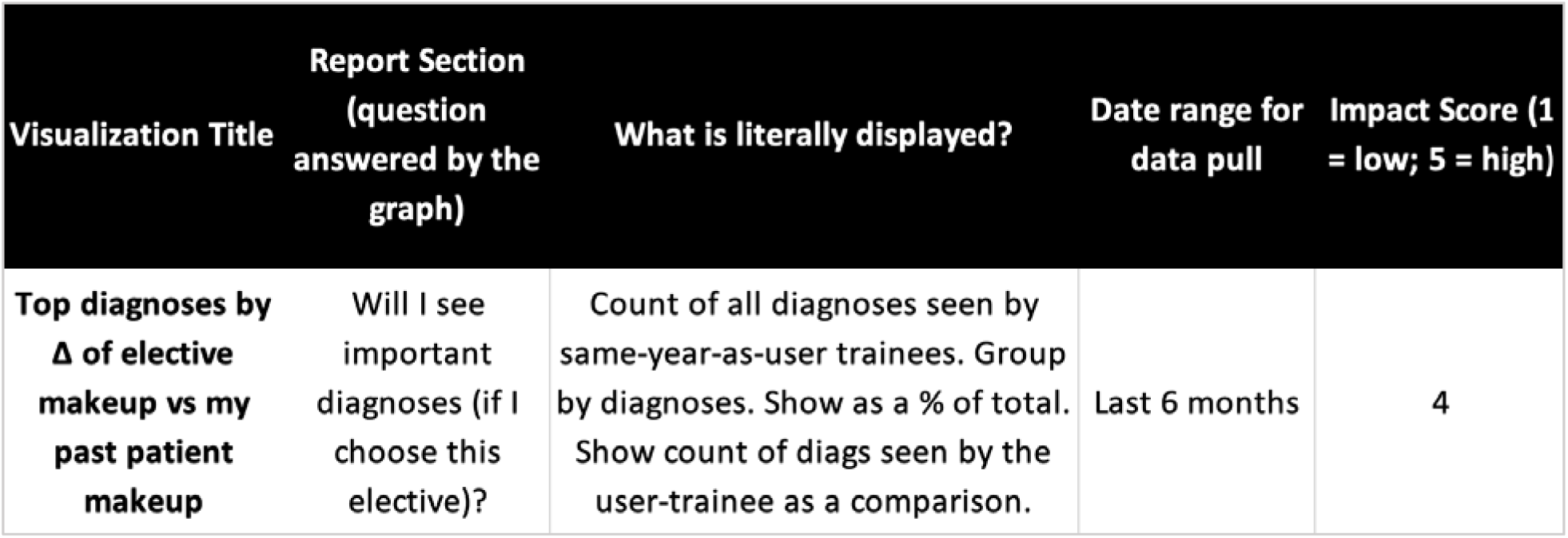
An example of a record, or single feature, from the feature prioritization matrix (FPM)

**Figure 2:**
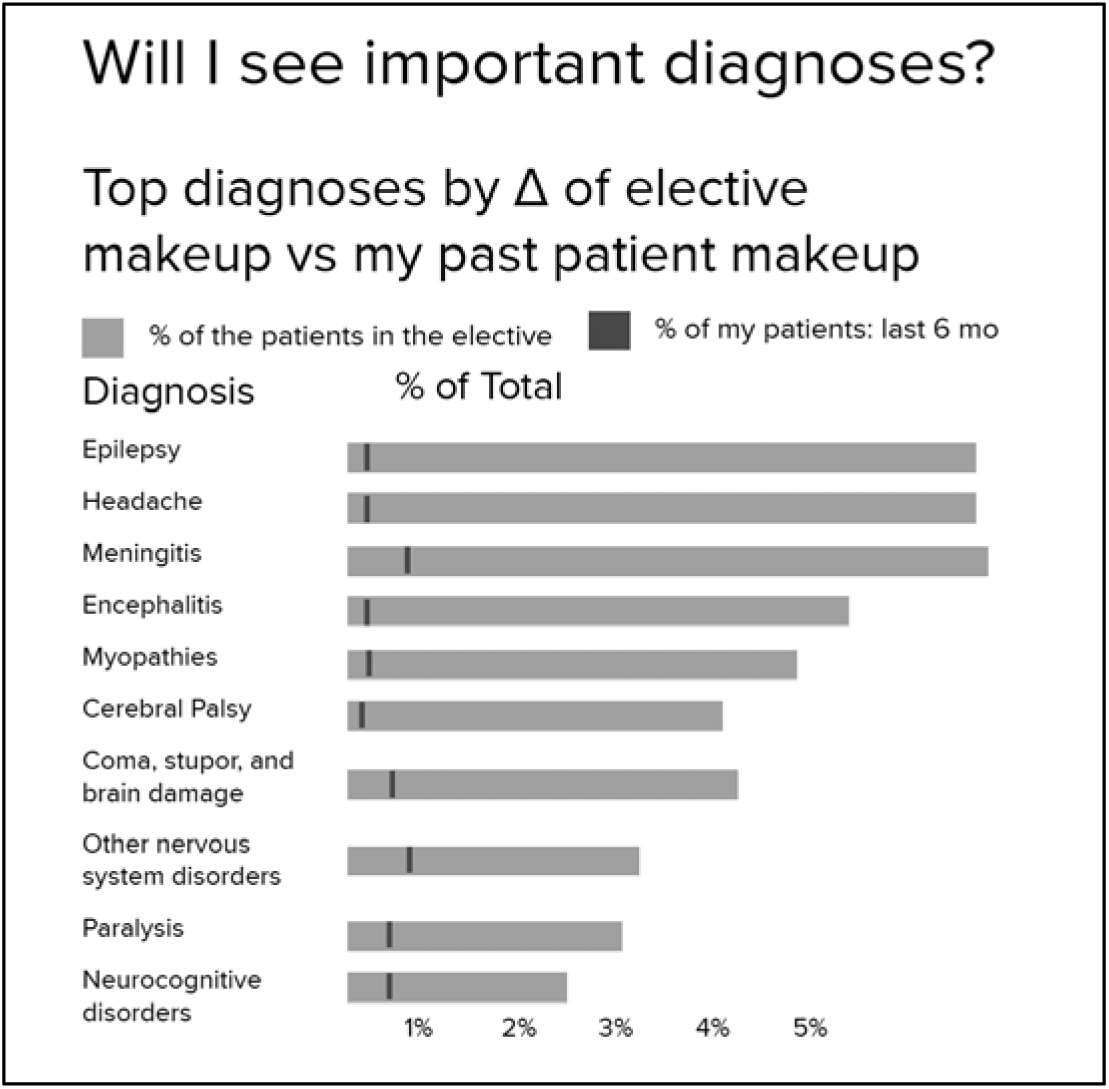
The corresponding visualization in the report from the feature described in Figure 1.

As we populated the FPM, the team began prototype development. We produced an initial midpoint report from a single collaborative unmoderated design session informed by reviewing recently completed process maps. We reached consensus on a set of visualizations that would advance planned learning, and these formed the first report. We iterated on the design in interview-walkthroughs with users (residents, fellows, and trainee mentors) until reaching saturation. With each walkthrough, newly elicited features and corresponding visualizations were added to the FPM and report. Once sessions repeated without eliciting new prototype feature categories, we considered saturation reached. We then produced a final “minimum viable” (MV) report in which the most impactful, feasible visualizations were included. Using the impact score and descriptions of required data for each visualization, we determined which features the current system architecture could feasibly support.

### Phase 3: Summative Usability Test

We developed a summative usability test to assess usability and utility of the MV midpoint report and measure performance on MAL planning related tasks using the report. There were five tasks, framed here as questions that residents ask themselves at key points in their experience. The MV report was meant to answer all of them.

1. What are the diagnoses to which I have received the most exposure?
2. In what clinical environments am I seeing them?
3. What gaps in diagnosis exposure could I be filling?
4. What is my exposure to acuity and complex care?
5. In each elective available to me, will I see important diagnoses?

For each task, we developed a scenario with realistic clinical data and a single complementary test question to be answered with an MV report feature. Four of the five scenarios were paired with a multiple-choice question. A fifth scenario was paired with a free-text response. Participants in this usability test were thus assessed with these five scenario-question pairs.

**Figure 3:**
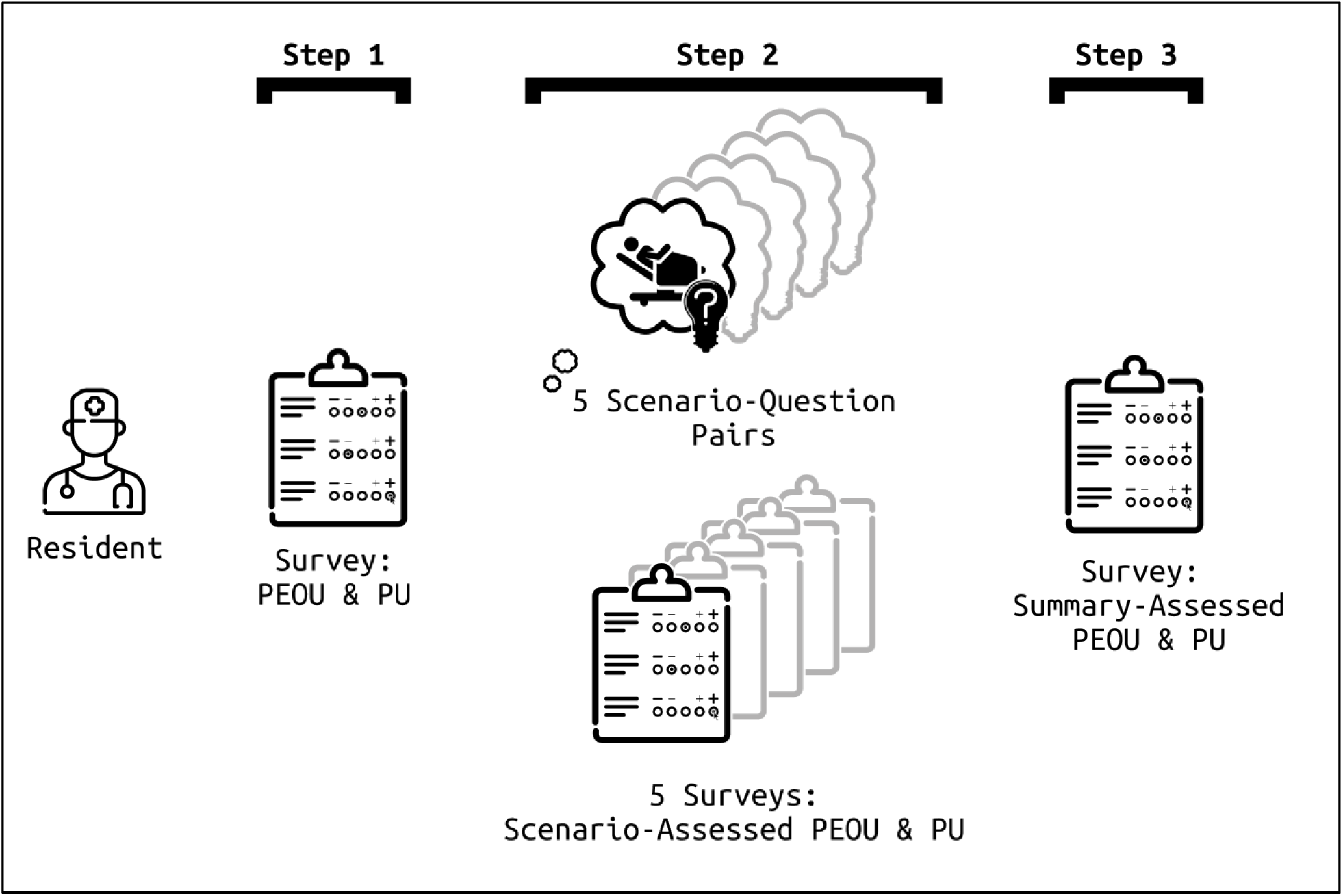
Summative UI test procedure for a single session, where a resident participant moves through the three steps, guided by a moderator.

We recruited a new cohort of resident trainees from the same pool of eligible subjects, excluding prior study participants. All data collection was performed over video call on Teams (Microsoft, Redmond WA) and Zoom (Zoom, San Jose CA). A single facilitator administered the test to each participant individually over the course of one hour. Participants were given the MV report in advance of the test for optional review. Participants were told to suspend disbelief and answer questions in the test as though the report were their own, aggregating their own practice area data and encountering the described scenarios during residency. Scenarios were presented via REDCap and we measured task times using time stamps in that system.

Participants completed a pretest questionnaire that asked PGY year of the participant, whether and how they kept records of their practice areas, and if so, perceived usability (PU) and perceived ease of use (PEOU) of that system. PU and PEOU questions were adapted from development of another successfully implemented CDS tool (Utidjian et al., 2015). Following each scenario-question pair, participants completed three PU and three PEOU ratings of the MV report using a nine-point Likert scale where responses at 6 – 9 were considered positive. Using the same scale, two questions in the post-scenario assessment addressed the realism of the PU assessment.

We report these descriptive statistics for the groups of measures that assessed PU and PEOU immediately after scenarios, PU and PEOU summarily at the very end of the test, and for the group assessing realism of the scenarios. Scenario-question pairs were scored for correctness and used to calculate task completion rate by dividing correct answers by total answered questions. We also report average task completion time as the amount of time spent from a scenario-question load page to the submission of a *correct* response.

## RESULTS

### Work Domain Assessment Results

We interviewed eight participants: one trainee, six program directors, and one administrator. We deployed initial descriptive and provisional coding efforts. Starting from the top of the codebook, 4 PETT scan categories decomposed into 34 topics which yet further broke down into 57 subtopics.

Through our process/causative coding and process map analysis we produced seven maps describing processes across three roles. The documents mapped 52 decisions or forks in the processes with 128 other steps, actions, or changes in mental model. Reviewing these maps, we determined that clinic and rotations were high-inertia settings with high mental stimulation. The Heavy demands on cognitive resources meant that planning in these situations would be poor or neglected completely. A report of practice area data in this environment would not be effective at supporting MAL if targeted for daily use.

**Figure 4:**
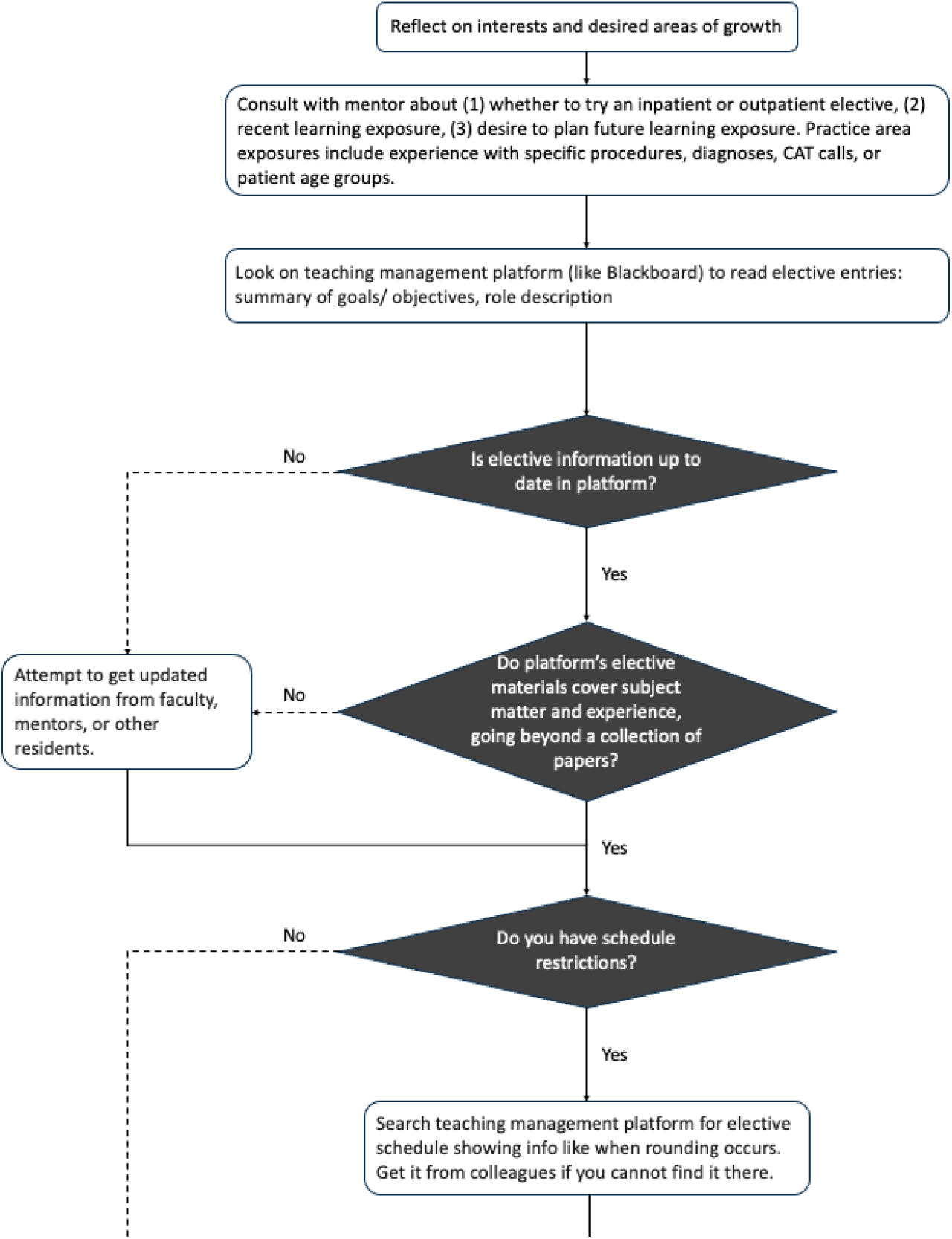
A segment of one of the seven process maps. Maps were simple, made with only two notation element-symbol pairs. Diamonds denoted decisions and rectangles stood for events, actions, or changes in mental model. Full process maps in sup materials.

Alternatively, trainee-mentor meetings, conducted much less frequently, stood out as a key target experience. By the time trainees meet with their mentors, they have usually formed some idea of a learning state gap but lack expert system knowledge required for planning its closure. These meetings offered a calm moment when trainees and mentors cooperatively analyzed a trainee’s learning states, both ideal and current, and planned to close the gap between the two. This trainee-mentor meeting environment had other benefits. It also allowed for contingency plans and provided a structure for re-planning in future meetings, both of which are optimal planning practices from the standpoint of human factors engineering and cognitive science.

**Figure 5:**
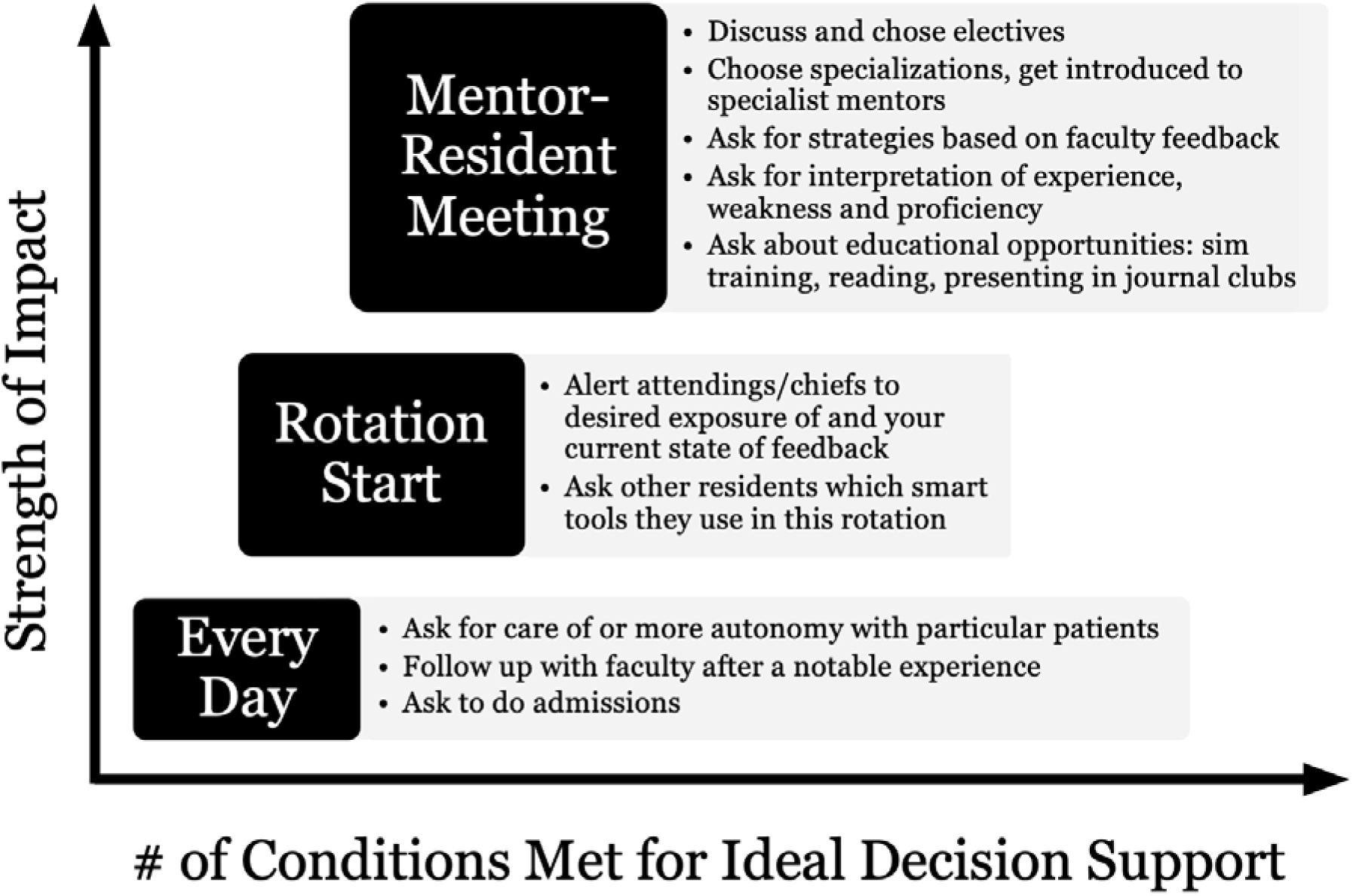
Settings where a report of practice area data could be leveraged and planned learning opportunities for each. These are plotted against qualities the research team used to select the target setting of the report.

### Formative Usability Test Results

Based on the results of our workflow assessment, we designed an initial report containing a single page of placeholder visualizations. Seven participants (six program directors and one fellow) completed the formative usability test, iterating from this initial version. Iterations to the midpoint report were made following each test, adding new and altering extant features. The version of the report following the final formative interview contained 51 distinct visualizations of trainee past and potential future data, spanning nine pages. The FPM contained a corresponding 51 records (see supplemental materials). Following feature reduction, the MV Midpoint report contained 5 visualizations over 3 pages, assuming one elective under consideration where an additional page would be added for each elective.

**Figure 6:**
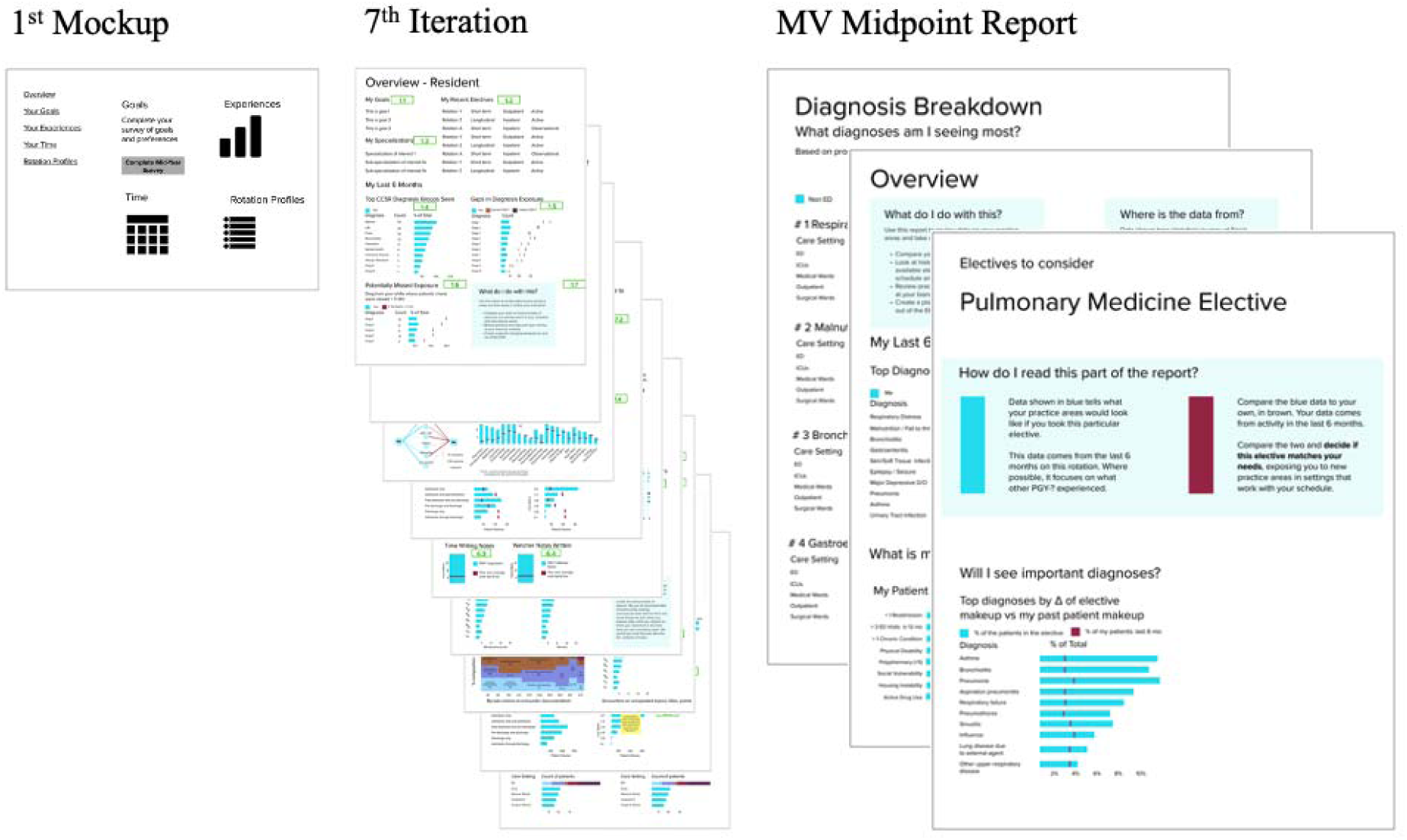
Progress of initial report to penultimate iteration to the final MV Midpoint Report used in Summative UI Test.

### Summative Usability Test Results

Eight resident physician participants completed the summative test. The task completion rate across all five scenario-question pairs, defined by the selection of the correct answer to a scenario’s accompanying multiple-choice question, was 78%. Mean task completion time was 2 minutes and 39 seconds (SD = 2:30).

Only one participant in the usability test reported keeping their own records on practice areas by “saving patient reports on Epic”, rating this method at a 5 in PU and PEOU, when scores 6-9 were considered positive.

| Measure | Score Group |
| --- | --- |
| This scenario is important in finding learning or entrustment gaps and resolving them. | Realism |
| This scenario is realistic in the search for learning or entrustment gaps and resolving them. | Realism |
| In this scenario, the system was easy to use. | PEOU |
| In this scenario, the system allowed me to perform tasks efficiently. | PEOU |
| Overall, I am satisfied with how the system is designed in this scenario. | PEOU |
| In this scenario, the system provided useful features. | PU |
| In this scenario, the system provided useful information. | PU |
| In this scenario, the system is an improvement over what I would have used before. | PU |

| Score Group | Median | IQR |
| --- | --- | --- |
| Scenario-Assessed PU | 8 | 4 |
| Scenario-Assessed PEOU | 8 | 3 |
| Realism | 8 | 3 |
| Summary PU | 8 | 1 |
| Summary PEOU | 7.5 | 1.25 |

The median score for Scenario-Assessed Perceived Usefulness was 8 (IQR = 4). Scenario-Assessed Median Perceived Ease of Use was 8 (IQR = 3). Scenario realism received a median score of 8 (IQR = 3). Distinct from the Scenario-Assessed PU and PEOU that followed each of the five scenarios, a single summary questionnaire assessed the same measures at the end of the test. Median Summary PU was 8 (IQR = 1). Median Summary PEOU was 7.5 (IQR = 1.25).

## DISCUSSION

In this study, we describe the human-centered design process used to develop a user interface to an educational decision support system. Like many designs for complex sociotechnical systems, an initial phase to better understand the implementation context led to substantial changes in our design goal. We shifted from a tool that trainees might use regularly to a tool for longer-term planning in the context of trainee-mentor meetings and potential choice of future elective rotations. We subsequently followed a typical iterative design and formative testing process. Upon achieving a stable “minimum viable” design, we conducted our summative testing. Given that our goal in summative testing is to predict future adoption of the tool by trainees, we supplement typical usability measures of task completion, efficiency, and perceived satisfaction with an assessment of perceived usefulness and intent to use, consistent with the Technology Acceptance Model (Holden & Karsh, 2010).

The ACGME requires trainees to experience a range of diagnoses during graduate medical training (ACGME, 2017). Using an education decision support system, we can facilitate appropriate diversity of patient experiences and better quantify outcomes in GME using informatics tools (Arora, 2018; Schumacher et al., 2019; Weinstein, 2017). Previous efforts to attribute trainee patient experiences have been limited in scope and dissemination. Several automated systems rely solely on EHR data (Levin & Hron, 2017; Schumacher et al., 2019). Yet these systems do not attribute patient experiences across clinical contexts or institutions. Dashboards to facilitate the feedback of clinical data have had mixed effectiveness, suggesting that learners – especially those in training – require facilitated precepting to understand their learning gaps, not just another dashboard (Hauer et al., 2018).

With the usability and perceived usefulness demonstrated in the summative test, we will implement the MV report to assist trainees with a number of tasks: visualizations of their top diagnosis types seen; their gaps in exposure compared to their peers and historical comparisons; their exposure to complex care patients; and most importantly, diagnosis makeup of electives or clinics that trainees could choose to further their exposure in weak areas of knowledge. The report will be delivered to trainees and their mentors before scheduled meetings biannually and will be discussed by the pair. Among other decisions that the report may inform, it will simplify the critical choice of electives or clinics that trainees can choose to better close their training gaps.

### Limitations

The study is limited by the study sample size at two institutions that may not represent the breadth of GME experience. Because of this, projected adoption based on high scores on PU and PEOU assessments may not be generalizable and ongoing measurement will be necessary during implementation.

The level of realism achieved in the simulations required participants to suspend disbelief. All participants had experience and context to create clear mental settings from experience in the medical education system, though fidelity achieved by descriptive text in immersing participants in that reality is low. In addition, the level of realism was tempered by technological limitations. We could not use actual trainee data from each specific participant or generalized trainee data from each site. Reports were populated by data that did not map to participants’ clinical experience, instead showing a generalized portrait of an “average” resident validated by clinical experts, requiring further suspension of disbelief.

## CONCLUSIONS

We iteratively developed and performed usability testing on a five-visualization report displaying trainees’ aggregate practice area data from the EHR. Our results indicate a high likelihood of the report’s adoption as an effective tool in graduate medical education, aligning with the MAL planning phase tasks and allowing them to be completed in a timely manner. We assessed use and usability with instruments and results predict significant future adoption. Evolving research will examine downstream effects on entrustment following implementation. This study provides a successful user interface for functions executed from an educational decision support system and offers a foundation for further research of clinical and educational applications of this system.

## Supporting information

Semi Structured Interview Guide for Nontrainees

Semi Structured Interview Guide for Trainees

Process Map - Mentor - Residency

Process Map - Program Director - Evaluations

Process Map - Program Director - Flow of Information to ACGME

Process Map - Program Director - New Policy Creation

Process Map - Resident - Elective Picking

Process Map - Resident - Mid Rotation

Process Map - Resident - Pre Rotation

Feature Prioritization Matrix

MV Midpoint Report

REDCap UI Test

## Data Availability

The datasets generated and/or analyzed during the current study are not publicly available due to data privacy restrictions established by our institutional review boards but are available from the corresponding author on reasonable request.

## List of Abbreviations

CDS: Clinical Decision Support
EDS: Educational Decision Support
EHR: Electronic Health Record
FPM: Feature Prioritization Matrix
GME: Graduate Medical Education
MAL: Master Adaptive Learner
MV: Minimum Viable
PETT: People, Environment, Tools, Tasks
PEUO: Perceived Ease of Use
PU: Perceived Usability
rPPI: Resident Provider Patient Interaction
SEIPS: Systems Engineering Initiative for Patient Safety
UCD: User Centered Design
UI: User Interface

## DECLARATIONS

### Ethics approval and consent to participate

The study was performed in compliance with the World Medical Association Declaration of Helsinki on Ethical Principles for Medical Research Involving Human Subjects. It was reviewed and approved by the institutional review boards at the University of Rochester (Study # 06717, Approved March 1, 2022) and the Children’s Hospital of Philadelphia (Study 21-018675 exemption granted August 8, 2021).

### Consent for publication

Not applicable

### Authors’ contributions

ACD and MVM conceptualized the study and designed the protocol along with NM and AF. AF, MVM, and ACD recruited subjects and conducted interviews. All authors participated in UCD process and analyzed interview results. AF conducted summative usability testing and prepared the draft manuscript. All authors read, edited, and approved the final manuscript.

## Acknowledgements

The authors would like to acknowledge the program leadership, administration, chief residents, and trainees in the pediatrics residency programs at the Children’s Hospital of Philadelphia and the University of Rochester.

## Authors’ Information

AF is a human factors engineer with experience in both user centered design for informatics and the healthcare industry. ACD and MVM are physicians and clinical informaticists with interests in using data-driven solutions to improve residency education. NM is a physician and clinical informaticist with a focus on cognitive informatics and decision support.

