## Supplementary material for "Precision Education Tools for Pediatrics Trainees: A Mixed-Methods Multi-Site Usability Assessment": Semi Structured Interview Guide for Nontrainees

Faculty Advisor and Mentor Interview Protocol

Note that all text in this format (blue, italicized) are notes to the interviewer/walkthrough facilitator. All unitalicized text in black is text which will be read aloud to the participant.

Participant ID:

We are developing a software tool to help residents be intentional about their education, giving them the chance to make more informed decisions. At the same time, it may equip mentors with information as well. The tool, named TRAILS, is a system that lets users access statistics drawn from the EHR, quantifying their experiences during residency. Most of the data in the Trails system come from clinician interactions with patients.

To make sure the system has the right features and is easy to use, we will ask about some of your meaningful experiences and decisions from your mentorship today.

We will also show you some sketches of the system, walk you through a few features it offers, and gather some reactions.

So, to outline again what we will be doing:

Asking you questions about experiences and decisions during when mentoring residents.

Gathering reactions to sketches of a tool.

All of the information that you provide will be kept confidential and your name will not be associated with your comments at any time. You are also able to stop at any time during the interview or not answer any questions you do not feel comfortable with. We would also like to ask that you try to avoid using identifiers by using pseudonyms where possible, but in case you do, know that when we transcribe the interview, that identifying data will be removed anyways.

- *Do you have any questions or concerns?*
- *Do you consent to participate in the study?*
- *Do you consent to being recorded?*

**Intro**

- Are you currently mentoring a resident?
- Does your mentorship come from a structured program or organization or was it informally requested?
- Can you list out the important stages in an internship?

**Interaction Types**

*In trainee mentorships, what are the interaction types, mediums, prompts, and patterns?*

- Tell us about the ways you interact with a current (or your most recent) mentee?
- What are the different events that would trigger a meeting, call, email, anything?
- What channels or mediums do you use to communicate (one on one, email, slack, etc)?
- Is there a pattern of types of interaction mapping to particular communication channels? For instance, do you use in-person meetings to have a specific type of discussion rather than email?
- Are there any significant mentor-meetings you remember?
  - First meeting?
  - First after end of internship? Etc?
  - Preceding a tough decision?
- Mentorship introduction meeting
  - How did you prepare?
  - How did the mentee prepare?
  - What were your goals for the meeting?
  - Did you reach all of those goals?
    - Why not?
  - How did you follow up on what was discussed?
- Standard meeting
  - How did you prepare for the most recent meeting with your mentee?
  - How did the mentee prepare?
  - What were your goals for the meeting?
  - Did you reach all those goals?
    - Why not?
  - How did you follow up on what was discussed?

**Information Gathering**

*What information gathering happens?*

- You listed several other interaction types earlier. *List them for the participant.* Tell us how you prepare for each of those.
- How did the mentee prepare for the recent meeting you described earlier?
- What are the ways that you currently learn about your mentee’s clinical experiences / the types and volumes of patients they have seen?
- What are you trying to learn when you go seeking information about their experience?
- Who do you reach out to?
- What obstacles are there to being effectively informed?

**Improvements**

*What defines mentorship success and failure?*

- Tell me about a mentorship that you thought was successful.
- How about a mentorship that was not successful.

**Feedback**

*What forms does feedback take, and what interactions contextualize it?*

- What are some of the ways mentees ask for your own feedback?
- a mentee’s reading plan?
- a mentee’s ILP or other goal writeup?
- a mentee’s duty hour logs?
- How long does it take you to respond to each type of feedback?
- Tell me about the last evaluation you filled out.
- How do you review faculty feedback with your mentee?
  - What are your goals in reviewing the feedback?
  - What are challenges on *your* end associated w those goals?
  - What are mentee challenges?
  - What are next steps?
  - What are challenges associated w those next steps?

**Micro and Macro Action to Improve Education**

*How have mentees taken action in the short and long term to improve comfort level and gain experience in practice areas?*

- Strategies
  - Talk about the balance of advising mentees to pursue experiences of interest and direct relevance to their future career path versus pursuing new, rounding-out, experiences that they won’t be able to get again after residency once dedicated to a path?
  - What informs advising them one way or another?
- Metacog
  - What are the ways that you ask mentees to be reflective?
  - Do you ask them to report back on their reflections?
  - What are the goals?
  - What are the barriers?
- Care Plan
  - How do you talk about growing care planning skills with your mentee?
  - Do you advise doing care plan comparisons between your mentee’s plans and attendings’ plans?
  - What traces of their thought process or reasoning can you find to look at their care planning skills?
- Can you think of a time when a mentee sought to or you advised a mentee to get more experience…
  - with a particular procedure (placing a line, lumbar puncture, anything)
    - For all questions in this list, probe on what prompted the decision and who had to be consulted or interacted with
  - with patients in a particular acuity level
  - with patients in a particular triage level
  - with patients in a particular age range
  - admitting patients
  - interpreting a particular test/diagnostic
  - interpreting a particular type of imaging
  - seeing higher overall volumes of patients per shift
  - with one of the 6 ACGME core competencies
- Think back to a time when a mentee evolved, going from a place of low comfort level in one of these practice areas to a place of high comfort level and autonomy. How did they get there?

**Information Gap**

*How have mentors seen and desired to see their mentees’ experience in residency quantified, on their own and compared to peer groups? What utility have they derived from it and what more could they gain?*

- Have you seen any of your mentees’ experience as a resident summarized and quantified?
  - [if so] What guidance did you give based on that data?
- Think about the measurable parts of residency we’ve covered so far. Imagine that you could quantify them and more, the entire residency experience. You can get every aggregated statistic about it. Imagine *also* that you could compare your mentee to their peers. You could do it at the rotation level, the elective, hospital-wide, resident year, national, any set of peers. Is that scenario clear?
  - Now imagine you’re looking through these stats, comparing how much experience your mentee has gotten in different measures to peers. What is the measure you look at and are most afraid to find them ranked low? What peer group level?
  - If you could show graphs of certain quantified experiences to your mentee, what would help you advise them most?
- What other information about your mentee’s performance would help you advise them better?

For below, when prototypes are available for demonstration, may proceed with this interview section.

**Prototype Demonstration**

Show the participant the prototype, walking through a maximum of three of the top expected interactions with the tool.

We are interested in a feature of the TRAILS system that we are calling the Midpoint Report. It would arrive, as a static PDF or packet midway through each year of training.

- What, if anything, did you like **least** about the system?
- What, if anything, did you like **most** about the system?
- What, if anything, was **missing** from the system?
- Additional comments:
