## Supplementary material for "Precision Education Tools for Pediatrics Trainees: A Mixed-Methods Multi-Site Usability Assessment": Semi Structured Interview Guide for Trainees

Interview Resident Protocol

Note that all text in this format (blue, italicized) are notes to the interviewer/walkthrough facilitator. All unitalicized text in black is text which will be read aloud to the participant.

Participant ID:

We are developing a software tool to help residents be intentional about their training and education, giving them the chance to make more informed decisions. The tool, named TRAILS, is a system that lets residents access statistics drawn from the EHR, quantifying their experiences during residency. Most of the data in the Trails system come from resident interactions with patients.

To make sure the system has the right features and is easy to use, we will ask about some of your meaningful experiences and decisions during residency today. We will also have an exercise for you to try to understand the mental model you have in place for those experiences and decisions. For that, we asked you to have some post-its or scraps of paper and a pen handy. Do you have those ready for later?

We will also show you some sketches of the system, walk you through a few features it offers, and gather some reactions.

So, to outline again what we will be doing:

Asking you questions about experiences and decisions during residency.

Doing an exercise to organize those ideas.

Gathering reactions to sketches of the tool.

All of the information that you provide will be kept confidential and your name will not be associated with your comments at any time. You are also able to stop at any time during the interview or not answer any questions you do not feel comfortable with. We would also like to ask that you try to avoid using identifiers by using pseudonyms where possible, but in case you do, know that when we transcribe the interview, that identifying data will be removed anyways.

- *Do you have any questions or concerns?*
- *Do you consent to participate in the study?*
- *Do you consent to be recorded?*

**Section for Interns Only**

*How and why have the intern’s habits around data logging of their experience and organization changed?*

- Have you ever kept your own log of data about the type or number of patients you see, different practice areas you perform, etc?
- If you know of another intern who does this, what do they track?
- Do you have a personal reference guide, for example adding information about high-yeild treatments or diagnoses, in Evernote or some other format?
  - If so Tell me about your most recent entry.
- Have you noticed other peers who are extremely efficient with their notes and workflows, leaving earlier while you are still working on signout notes?
  - If so Tell me what seeing that has triggered in reflection or change on your part?
- Do you use dot phrases?
  - If so What prompted you to start?
  - If not Why not?
- Do you generally use note templates?
  - If so Do you specifically use note templates for your most common diagnoses?
  - If not Why not?
- Would you want to know your most commonly typed words/phrases in notes?

**Macro Action to Improve Education: Rotation and day to day**

*What behaviors and interactions create an environment for learning and mediate intentional learning at the rotation level?*

- When you start a new rotation, how do you prepare for it?
- Have you ever attempted to learn the historical makeup of patients that present in a particular rotation, for example, medical history, SES, treatment compliance, prior visits to CHOP, or really anything else in aggregate?
  - Tell me about the most recent example.
  - Where did you get the data (system name, how accessed)
  - Who did you deal with to get the information (role, how contacted)
  - If not Why not?
- How have you tried to place yourself “at the right place at the right time”, for example, assisting with an after-hours case or making yourself available for added duties?
- Do you prepare for rounds at the start of a new rotation differently than at the end of a rotation?
- Have you ever made an organized effort to note and learn the names of new nurses and techs at the start of a new rotation?
- Resident - Getting experience with admissions
  - How do you/could you, as a resident, advocate for attending permission to do admissions and get practice?
  - Do you show them shared docs about who’s done them?
  - What else happens?

**Macro Action to Improve Education: Electives**

*What motives, interactions, obstacles, and frustrations characterize the decision process around and enrollment in electives, with particular focus on information and individuals?*

- Tell us about the last elective you decided to take. Tell us specifically about what led to that decision.
- Did you ever make a difficult decision between two electives? What made you choose one over the other?
- Did you have to convince anyone to let you enroll? Who and how?
- Was there a time when you could not get the elective you wanted to take?
- Were there things you wanted to know about the elective that you could not find out before you were required to decide?
- If you could have any stat about the experience other residents had in that elective or that physicians in general experience in that department, what would you want to know?
- If you could design your own elective, what would that look like? What information would you need to better design it
- Resident - Getting experience with admissions
  - How do you/could you, as a resident, advocate for attending permission to do admissions and get practice?
  - Do you show them shared docs about who's done them?
  - What else happens?

**Macro Action to Improve Education: Mentorship and Feedback**

*What informs and what results flow from impactful mentorship?*

- Tell me about someone who has effectively guided you in residency.
  - Was this person a formal mentor?
  - Where did they get information about your experience to ground their guidance?
  - Walk me through a piece of advice and an action you took as a result from them.
- Tell me about the last evaluation faculty filled out for you on the 6 ACGME core competencies.
  - What actions did you take as a result?
- Describe your mental model of how mentorship works?
- What are the target outcomes/goals of mentorship?
- What does an effective mentorship relationship look like?
- Are you doing care plan comparison between your plan and attending’s plan and reporting back to your mentor?
- Were you advised to do so? Anything similar?
- Are mentors advising you towards metacog/reflective/report-back strategies?
- Are they valuable when you do them if done at all?
- Are you encouraged to stick with what you like or try many new things during residency?
- What do you talk about to mentors (and directors) when you talk about your wellbeing?
- Walk me through the important stages in mentorship?
- Are there any significant mentor-meetings you remember?
  - First meeting?
  - First after end of internship? Etc?
  - Preceding a tough decision?
- Mentorship introduction meeting
  - How did you prepare?
  - What were your goals for the meeting?
  - Did you reach all of those goals?
    - Why not?
  - How did you follow up on what was discussed?
- Standard meeting
  - How did you prepare?
  - What were your goals for the meeting?
  - Did you reach all those goals?
    - Why not?
  - How did you follow up on what was discussed?
- Strategies
  - Are you encouraged to stick with what you like or try many new things during residency?
- Metacog
  - What are the ways that mentors ask you to be reflective?
  - Do they ask you to send them results of your reflection?
  - Are they valuable?
- Care Plan
  - How do you talk about growing care planning skills with your mentor?
  - Are you doing care plan comparison between your plan and attending’s plan and reporting back to your mentor?
  - Were you advised to do so? Anything similar?
- Wellness/Own Mental Health
  - What do you talk about to mentors (and directors) when you talk about your wellbeing?
- Faculty Feedback
  - How do you review this with your mentor?
  - What are your goals in reviewing the feedback?
  - What are challenges associated w those goals?
  - What are next steps?
  - What are challenges associated w those next steps?

**Fellowship Application**

*What data collection and preparation on practice areas and aggregated patient data from their residencies do residents intend to and wish they could perform for use in fellowship applications?*

- Do you plan to do a fellowship? If “no”, skip this section.
- What information about your whole resident experience will you try to gather to prepare an application for fellowship?
- If you could wave a wand and get any stats about your time in residency, what would you want to see?
- If you were going to include one graph to represent your time in residency on your fellowship application, what would be on it? Can you sketch it out for me now?

**Conferences**

*What data collection and preparation on practice areas and aggregated patient data from their residencies do residents intend to and wish they could perform for use in conferences?*

- If you need to dig into cases you have seen to prepare for a medical conference, describe that process?

**Micro Action to Improve Education**

*What are residents’ experiences seeking more experience in specific practice areas, growing in comfort levels, and autonomy and what motivated them?*

- Tell us about a time when you sought to get more experience…
  - with a particular procedure (placing a line, lumbar puncture, anything)
    - For all questions in this list, probe on what prompted the decision and who had to be consulted or interacted with
  - with patients in a particular acuity level
  - with patients in a particular triage level
  - with patients in a particular age range
  - admitting patients
  - interpreting a particular test/diagnostic
  - interpreting a particular type of imaging
  - seeing higher overall volumes of patients per shift
  - with one of the 6 ACGME core competencies
- So this far, we’ve asked about some of the levers that you can pull as a resident to go from a place of low comfort level in one of these practice areas to a place of high comfort level and autonomy…. List some of those mentioned. What other levers can you pull to take your education into your own hands?
- Think back to a recent patient interaction when you were front lining autonomously, doing something that, as an intern, you would be constantly checking in with the chief resident or attending. Tell me about the journey to get there.

**Information Gap**

*How have residents seen and desired to see their experience in residency quantified, on their own and compared to peer groups? What utility have they derived from it and what more could they gain?*

- Have you seen any of your experience as a resident summarized and quantified?
  - if so…
    - What immediate changes happened because you got that information? If unclear, clarify that you’re looking for actions they took or environmental changes that happened.
    - What long term changes happened?
    - Do you remember *feeling* differently about your experience because of that information?
- Think about the measurable parts of residency we’ve covered so far. Imagine that you could quantify them and more, your entire residency experience. You can get every aggregated statistic about it. Imagine *also* that you could compare yourself to your peers. You could do it at the rotation level, the elective, hospital-wide, resident year, national, any set of peers. Is that scenario clear?
  - Now imagine you’re looking through these stats, comparing how much experience you’ve gotten in different measures to peers. What is the measure you look at and are most afraid to be ranked low at in experience? What peer group level?
  - If you could show graphs of certain quantified experiences to your mentor, on what would you most want feedback?
  - What would you pull up to fill out your ILP?
  - What would you pull up when logging information for ACGME (procedures?)?
  - What would you pull up to learn about and improve your efficiency in day to day work?
- Compared to you peers, would you want to know…
  - how frequently you checked in on your average patient’s chart
  - how frequently you went to check on orders on average
- you arrived on time

**Prototype Demonstration**

If available, show the participant the prototype, walking through a maximum of three of the top expected interactions with the tool.

- What, if anything, did you like **least** about the system?
- What, if anything, did you like **most** about the system?
- What, if anything, was **missing** from the system?
- Additional comments:
