## Supplementary figures and images for "Precision Education Tools for Pediatrics Trainees: A Mixed-Methods Multi-Site Usability Assessment"

### Process Map - Mentor - Residency

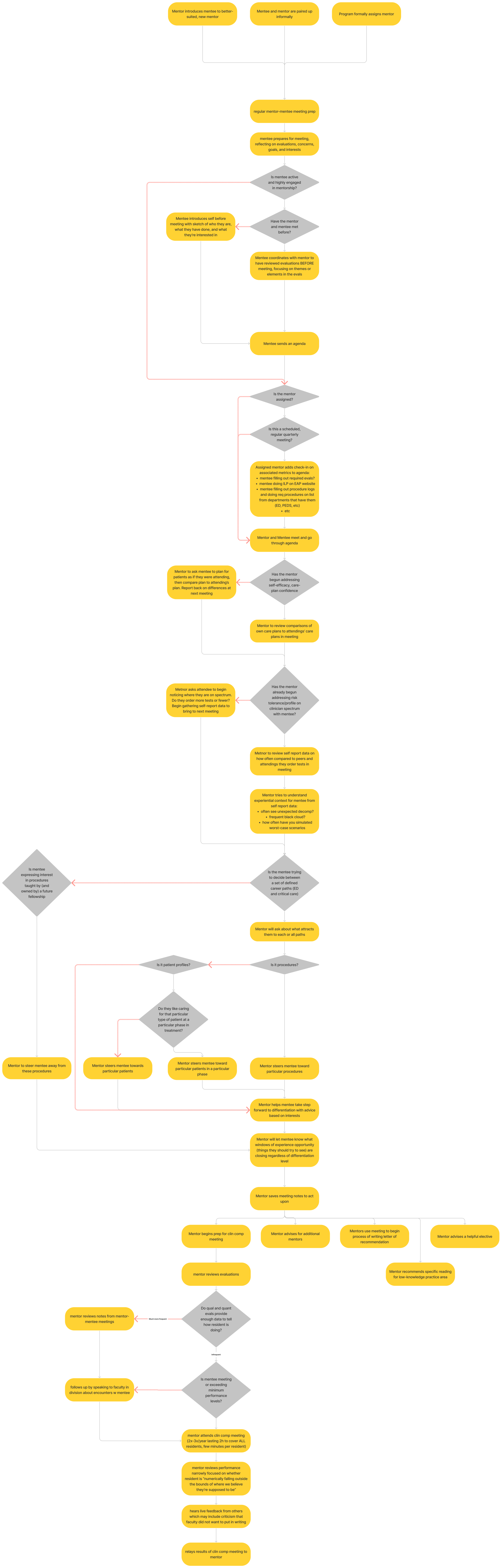

### Process Map - Program Director - Evaluations

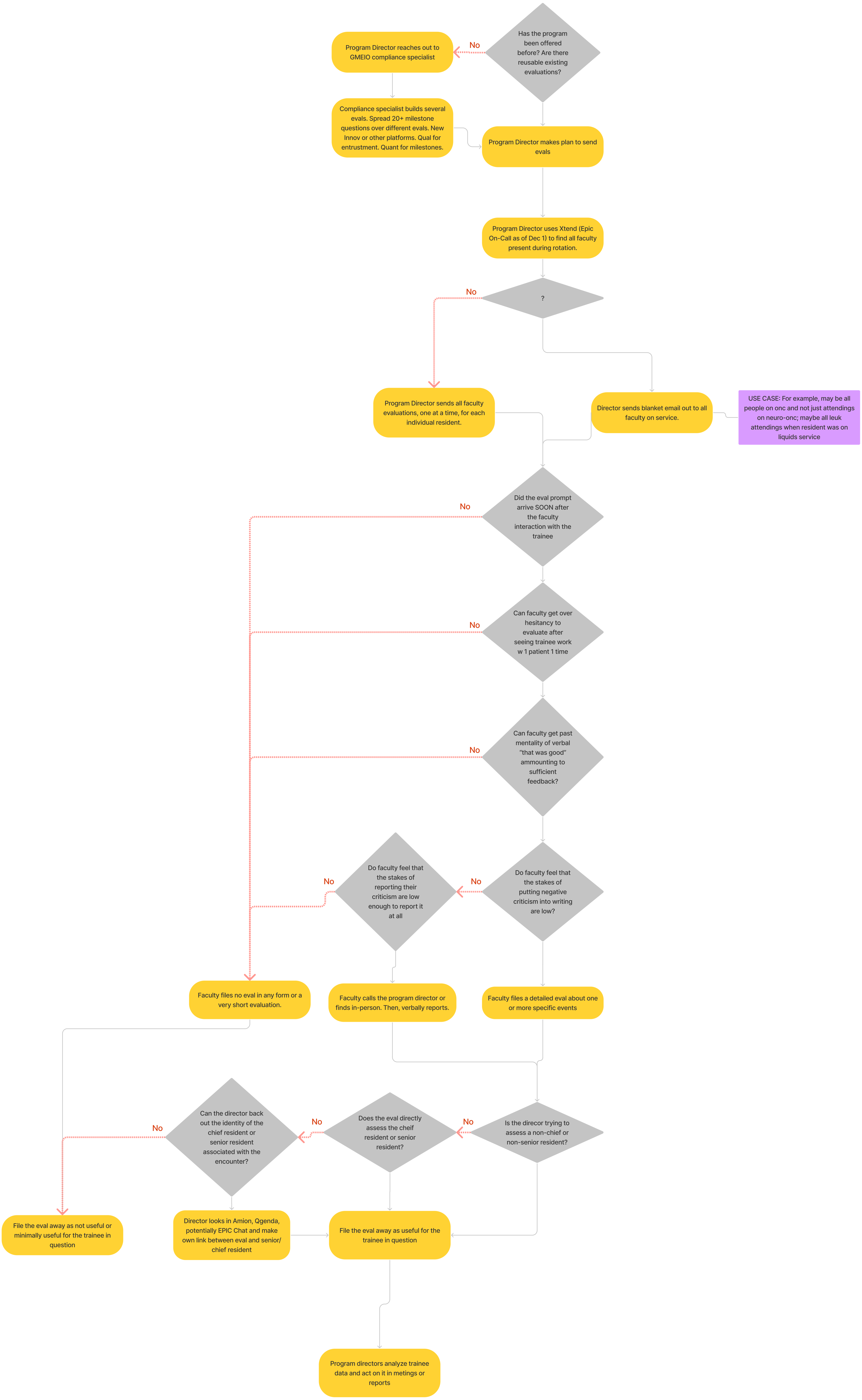

### Process Map - Program Director - Flow of Information to ACGME

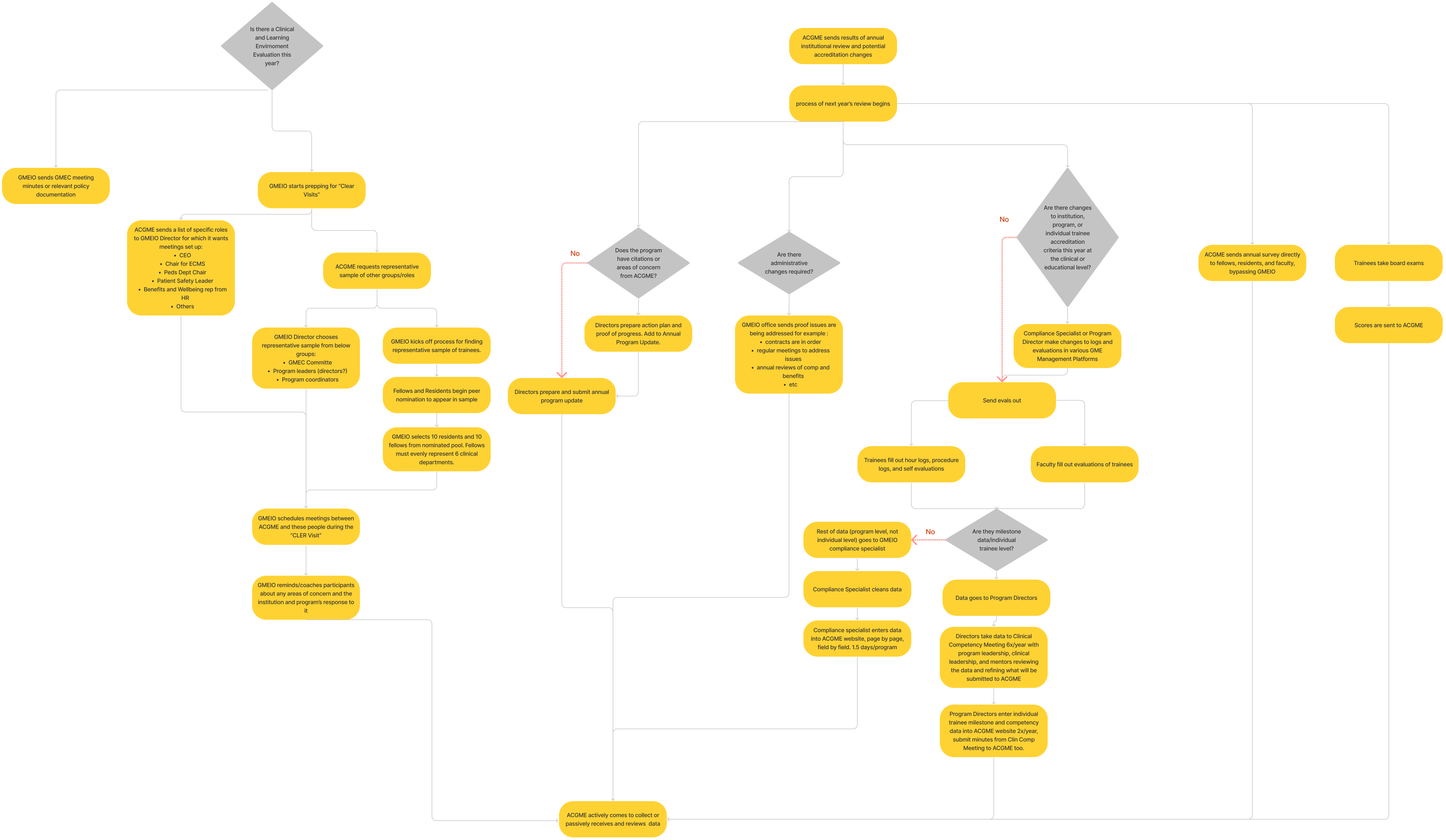

### Process Map - Program Director - New Policy Creation

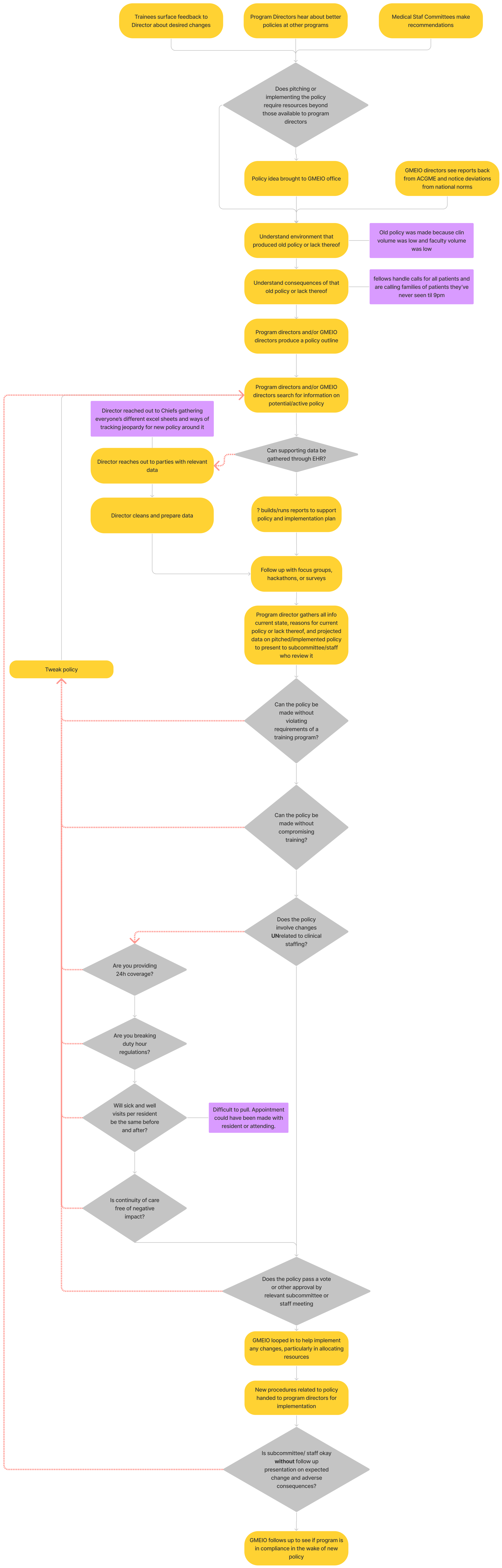

### Process Map - Resident - Elective Picking

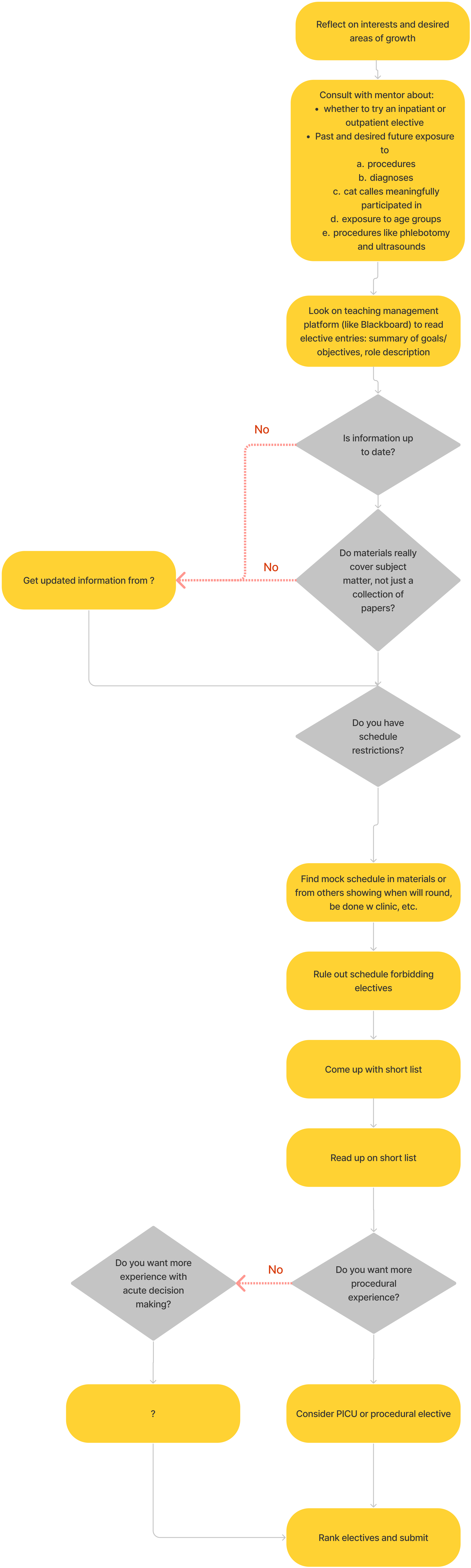

### Process Map - Resident - Mid Rotation

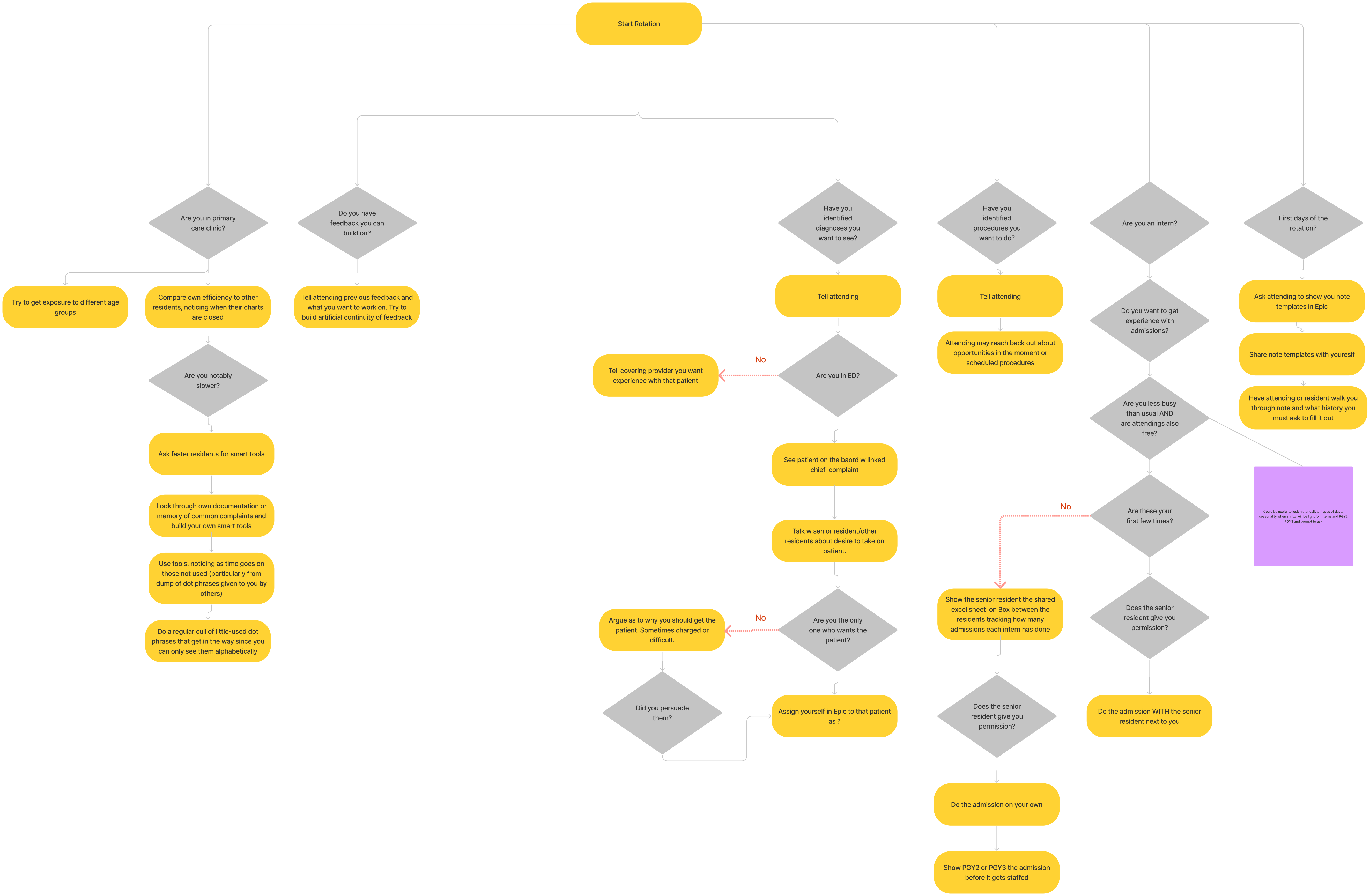

### Process Map - Resident - Pre Rotation

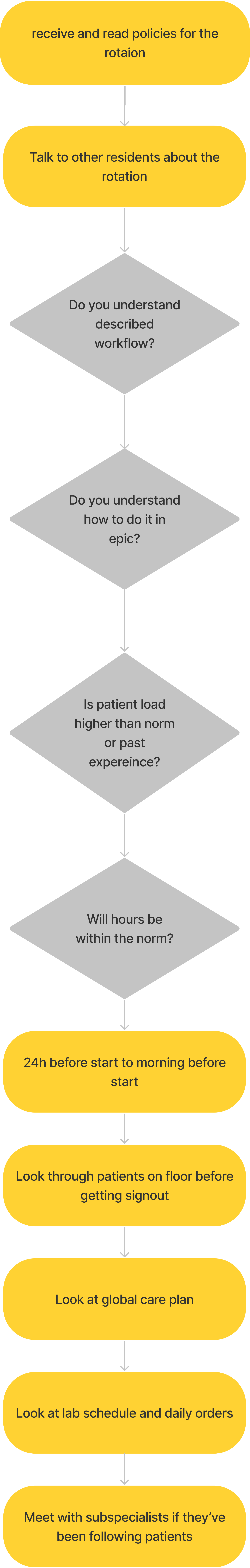
