## Supplementary material for "Precision Education Tools for Pediatrics Trainees: A Mixed-Methods Multi-Site Usability Assessment": Feature Prioritization Matrix

Each record (a row in the matrix with an entry for each of those 3 data types) had a corresponding data visualization in the midpoint report. The visualization showed a report owner's aggregate data for some element of their clinical experience or forecast potential future exposures (e.g., how many patients with a particular diagnosis had they seen in the last six months set against how many they might see on a particular elective). The data for each visualization was described in the form of the structured query required to generate the visualization, written adapted to more natural language.

| Is it V1, V2, or V3? | Visualization Title | Section (question answered by the graph) | What is literally displayed? | Date range for data pull | Impact Score (1 = low; 5 = high) |
|---|---|---|---|---|---|
| V1 | Top CCSR Diagnosis Group Seen | My Last 6 Months | Count of diagnoses from user trainee's RPPI; Count of total patients that had an RPPI with user trainee. Group by CCSR diagnosis type. Show the top 10 diags by volume. Show count and show diag count as fraction of total patients. | Last 6 months | 4 |
| V1 | Gaps in Diagnosis Exposure | My Last 6 Months | Count of diags from user trainee's RPPI. Count of diags for all same-year-as-user trainees. Count of diags from all trainees over past years (flexible number based on how hard this is to pull; 2-5 years maybe?). Group the counts by diag and by whether they belong to the user trainee, their current cohort, or past years. Calc the difference between current cohort diag counts and user-trainee's diag count. Sort by the difference, descending. Show the top 10 diags by difference. Show counts for user trainee, cohort, and historic cohort where hist cohort divided by (total months of pull/6). | Last 6 months | 4 |
| V1 | What diagnoses am I seeing Most? | Diagnosis Breakdown | Count of diags. Group by care setting (ED, ICUs, med wards, outpatient, and surgical) and by acuity level (when the care setting is the emergency department). | Last 6 months | 4 |
| V1 | Top diagnoses by Δ of elective makeup vs my past patient makeup | Will I see important diagnoses (if I choose this elective)? | Count of all diags seen by same-year-as-user trainees. Group by diags. Show as a % of total. Show count of diags seen by the user-trainee as a comparison. | Last 6 months | 4 |

| | | | | | |
|---|---|---|---|---|---|
| V1 | My Patient Makeup | What is my exposure to acuity and complex care? | Count of all user trainee's RPPI patients. Of those, fraction that had more than one readmission; fraction that had more than 3 emergency department visits in 12 months; fraction that had more than one chronic condition; fraction that had more than 5 medications; that suffer housing instability; that are active drug users. | Last 12 Months | 3 |
| V1 | (no title - explainer on where data comes from) | Readme | Note about where the data comes from | na | 3 |
| V1 | What do I do with this? | Readme | Explains how to use the report | Last 6 months | 2 |
| V1 | How do I read this part of the report | Readme | Note about how to read and use the report | na | 2 |
| V2 | Potentially Missed Exposure | My Last 6 Months | Count of all diags from RPPI with the user trainee. Sum of time user trainee opened charts of any patients seen by any clinician on the user trainee's shifts where. Where sum of time in those charts is <5minutes, treat that patient as a missed exposure. Count all diags from missed exposures. For all diags that makeup bottom 80% of actual exposure, show the top diags by missed exposure. Sum actual exposure to missed exposure counts as Potential Exposure. Group by diag. Show actual exposure by count and % of total exposure and show Potential exposure. | Last 6 months | 4 |
| V2 | What is the time commitment | What is the time commitment? | time logged in the EHR from different terminals and devices: Mobile, Trainee Home machine, and CHOP machines | Last 6 months | 4 |
| V2 | Diagnoses my mentors want me to see (elective/clinic comparison) | Will I see important diagnoses? | Same as 7.1 but filtered on diags that fall into CCSR types that were selected by mentors as a target set for trainees to get experience with. | Last 6 months | 4 |
| V2 | Residents who took this elective | Who can I ask about it? | show other trainees who have taken the elective/clinic | Last 2 years | 4 |
| V2 | My Specializations | Overview (pre-graphs) | Short set of specialization interests (no more than 3) entered in by the user following a prompt when they first log into the system. Should be low character limit in the field entry form. | Last 6 months | 3 |
| V2 | Time Writing Notes | How active and demanding is it? | time spent writing any type of notes in an elective | Last 6 months | 3 |

| | | | | | |
|---|---|---|---|---|---|
| V2 | Continuity of Care (elective/clinic comparison) | How much exposure will I get to each patient? | number of RPPI average patients for same-year-as-user trainees at each level of a continuity of care metric (COCM). Two ideas for COCM metrics: [1] the validated Usual Provider of Care (UPC - see cited paper) metric [2] number of sequential visits to or encounters with each trainee. Group by COCM rounded to nearest whole number. Show data from graph 3.9 compared to same-year-as-user trainees data. | Last 6 months | 3 |
| V2 | Rotation's Patient Makeup | Will I see complex care? | Same as 3.7 but for all same-year-as-user trainees. | Last 6 months | 3 |
| V2 | Concurrent EHR Sessions Per Day on the Same Patient | Will I coordinate care with other roles? | See reference articles for calculating metric. | Last 6 months | 3 |
| V2 | Continuity of Care | How much exposure am I getting to each patient? | Count of of RPPI patients for user trainee at each level of a continuity of care metric (COCM). Two ideas for COCM metrics: [1] the validated Usual Provider of Care (UPC - see cited paper) metric [2] number of sequential visits to or encounters with user trainee. Group by COCM rounded to nearest whole number. | Last 6 months | 2 |
| V2 | New Patients | How much exposure am I getting to each patient? | Count of patients where patients had no prior history with the Unit or clinic and an RPPI with the user trainee is their first. | Last 6 months | 2 |
| V2 | Attention Switches | How efficient and acurate am I with the EHR? | See reference articles for calculating attention switches. | Last 12 Months | 1 |
| V3 | What diagnoses do my mentors want me to see? | Diagnosis Breakdown | Same as 2.1 but filtered on diags that fall into CCSR types that were selected by mentors as a target set for trainees to get experience with. | Last 6 months | 4 |
| V3 | My Goals | Overview (pre-graphs) | Short set of goals (no more than 3) entered in by the user following a prompt when they first log into the system. Should be low character limit in the field entry form. | Last 6 months | 3 |
| V3 | My Recent Electives | Overview (pre-graphs) | Pull list of electives in last 6 months | Last 6 months | 3 |

| | | | | | |
|---|---|---|---|---|---|
| V3 | CAT Notes | What is my exposure to acuity and complex care? | Count of CAT notes written by the user trainee OR written about any RPPI patient of the user trainee's within +/- 24h that the user trainee saw them. Group by month. | Last 12 Months for raw data; filter on Hour +/- 24h of user trainee's shifts | 3 |
| V3 | Watcher Notes | What is my exposure to acuity and complex care? | Count of Watcher notes written by the user trainee OR written about any RPPI patient of the user trainee's within +/- 24h that the user trainee saw them. Group by month. | Last 12 Months for raw data; filter on Hour +/- 24h of user trainee's shifts | 3 |
| V3 | Any of the above | What is my exposure to acuity and complex care? | Sum patient volumes from 3.1 , 3.2, 3.3, and 3.4. Group by month. | Last 12 Months for raw data; filter on Hour +/- 24h of user trainee's shifts | 3 |
| V3 | Time reading ICU consult notes | What is my exposure to acuity and complex care? | Sum of user trainee time spent reading notes where notes are written by ICU consults. | Last 6 months | 3 |
| V3 | Sample Quality by BX Type x Imaging | What is my exposure to procedures? | Average of sample quality scores from Procedure Master (/Model?) File for all biopsies where user trainee performed the biopsy. Compare to same but performed by attengings only during user trainee's shifts. Should show if the samples fellows are getting are actually good samples? Are the quality scores of the samples from radiology improving? | Last 6 months | 3 |
| V3 | Time reading consult notes by dept | Do I see coordinated care? | Sum of time for all same-year-as-user trainees spent reading notes where notes are written by consults. Group by consult's department. Get average time/week for user trainee. | Last 6 months | 3 |
| V3 | Acute Patient Volume | Will I see acuity? | Count of all patients with acute criteria (CAT notes, Watcher notes, ICU Transfers, or patients put on ventilator or pressors) in the elective. Get weekly average. Compare to sum from graph 3.5 made into weekly average. | Last 6 months | 3 |

| | | | | | |
|---|---|---|---|---|---|
| V3 | Time Reading Consult Notes (elective/clinic comparison) | Will I see acuity? | Sum of time for all same-year-as-user trainees spent reading notes where notes are written by consults. Group by consult's care setting (ICU, surgical ward, medical ward). Get average time/week for rest of class vs trainee. | Last 6 months | 3 |
| V3 | Patients By Phase In Their Hospital Course | How much exposure will I get to each patient? | Patient volume where same-year-as-trainees were the provider during any of the following buckets: (1) Admission ONLY (2) Admission AND any other care before discharge (3) any care between discharge AND NOT Admission AND NOT Discharge (4) any care before discharge AND discharge (5) Discharge ONLY (6) Admission AND any care AND discharge. Compare to user trainee's stats for each bucket from 3.8 | Last 6 months | 3 |
| V3 | Time reading consult notes by dept (elective/clinic comparison) | Will I coordinate care with other roles? | Same as 5.1 but instead, pull for all same-year-as-user trainees in the elective/clinic. | Last 6 months | 3 |
| V3 | Transferred to ICU | What is my exposure to acuity and complex care? | Count of transfers to ICU of any RPPI patient of the user trainee's within +/- 24h that the user trainee saw them. Group by month. | Last 12 Months for raw data; filter on Hour +/- 24h of user trainee's shifts | 2 |
| V3 | Hospital Course Exposure | How much exposure am I getting to each patient? | Count of patients where user Trainee was the provider during any of the following buckets: (1) Admission ONLY (2) Admission AND any other care before discharge (3) any care between discharge AND NOT Admission AND NOT Discharge (4) any care before discharge AND discharge (5) Discharge ONLY (6) Admission AND any care AND discharge | Last 6 months | 2 |
| V3 | Procedures Performed by Month | What is my exposure to procedures? | Count of procedures where user trainee led or assisted the procedure or where an attending performed the procedure when the trainne was on shift. Group by user trainee involvement vs not (shown as attending's procedure) and by month. | Last 12 Months | 2 |
| V3 | By Care Setting | What is my exposure to procedures? | Same as 4.1 but instead, second Group By is care setting (ICU/Surgical/Medical) | Last 6 months | 2 |

| | | | | | |
|---|---|---|---|---|---|
| V3 | By Procedure Type | What is my exposure to procedures? | Same as 4.1 but instead, second Group By is CCSR procedure type (major/minor x theraputic/diagnostic). Map from ICD 10 codes here: https://www.hcup-us.ahrq.gov/toolssoftware/ccsr/prccsr.jsp | Last 6 months | 2 |
| V3 | By Procedure | What is my exposure to procedures? | Same as 4.1 but instead, second Group By is is the procedure itself | Last 6 months | 2 |
| V3 | Education by Method and Response to it | Is my patient education skillset improving? | Count of "Method" and "Response" values in the Epic Education Topic data entry form. Group by method, then by response. Response should be shown as a percent of all responses in that particular method. For example, when the method is "Handout" show the percent of all Handout encounters for each of the five response values. | Last 6 months | 2 |
| V3 | Missed opportunities to repeat existing education | Is my patient education skillset improving? | Count of RPPI where either of the two preceding encounters (regardless of the clinician involved) included new education AND the user trainee (during their RPPI) did not repeat/reinforce the topic. | Last 12 Months | 2 |
| V3 | Watcher Notes Written | How active and demanding is it? | time spent writing ONLY watcher notes | Last 6 months | 2 |
| V3 | Phone Calls to Families | How active and demanding is it? | Number of phone call notes or calls logged otherwise in EHR | Last 6 months | 2 |
| V3 | What procedures will I see? | What procedures will I see? | Count of procedures where same-year-as-trainee population performed the procedures and procedures where up-authority personnel performed the procedure and not the trainee. For a resident trainee, also count procedures performed by fellows and attendings. For a fellow trainee, also count the attending procedures. Group by trainee performed procedures, by up-authority performed procedures, and by procedure itself. | Last 6 months | 2 |
| V3 | Education Method Volume | Will my education skillset grow? | Similar raw data to 4.5<br>Count of encounters for all same-year-as-user trainees where education topic data was entered. Group by education method. Compare to count of encounters for the same thing for User trainee. Show average by week. | Last 6 months | 2 |

| | | | | | |
|---|---|---|---|---|---|
| V3 | Education Response | Will my education skillset grow? | Similar raw data to 4.5<br>Count of encounters for all same-year-as-user trainees where education topic data was entered. Group by response to education. Compare to count of encounters for the same thing for User trainee. Show average by week. | Last 6 months | 2 |
| V3 | Education Repetition Opportunities | Will my education skillset grow? | Similar to 4.6<br>Count of RPPI in the elective/clinic where either of the two preceding encounters (regardless of the clinician involved) included new education (implying that counted encounter got the chance to repeat/reinforce the topic). Compare to same value for user trainee. | Last 6 months | 2 |
| V3 | Total Labs and Imaging Ordered | Will it be a diagnostic stewardship and interpretation challaenge? | For all same-year-as-user trainees, number of weekly labs and images ordered in the elective/clinic. Compare to user trainee's max and minimum orders of the same diagnostics for all weeks. | Last 6 months | 2 |
| V3 | Top 10 Diagnostic Orders | Will it be a diagnostic stewardship and interpretation challaenge? | Same as 9.1 but grouped by Diagnostic. | Last 6 months | 2 |
| V3 | Put on Vent or Pressors | What is my exposure to acuity and complex care? | Count of user trainee's RPPI patients who were put on a ventilator or vasopressors within +/- 24h that the user trainee saw them. Group by month. | Last 12 Months for raw data; filter on Hour +/- 24h of user trainee's shifts | 1 |
| V3 | Medication Errors | How efficient and acurate am I with the EHR? | See reference articles for calculating metric. | Last 12 Months | 1 |
| V3 | Wrong-Patient Errors | How efficient and acurate am I with the EHR? | See reference articles for calculating metric. | Last 12 Months | 1 |
| V3 | Ambulatory EHR Efficiency | How efficient and acurate am I with the EHR? | See reference articles for calculating metric. | Last 12 Months | 1 |
| V3 | Time writing messages that are opened for >1 second or not at all | How efficient and acurate am I with the EHR? | See reference articles for calculating metric. | Last 12 Months | 1 |
