## Supplementary material for "Precision Education Tools for Pediatrics Trainees: A Mixed-Methods Multi-Site Usability Assessment": MV Midpoint Report

### Overview

#### What do I do with this?

Use this report to review data on your practice areas and take steps to further your education.

- Compare your stats to cohort averages.
- Look at historical data from residents in available electives and decide which fit your schedule and educational needs
- Review practice area data with your mentor at your biannual meeting
- Create a plan for changing behaviors in and out of the EHR

#### Where is the data from?

Data shown here start their journey at Epic's Clarity relational database that records all interaction with the software. We execute a series of scripts that extract fields of interest. We pull all structured data clinicians enter, leaving unstructured data behind. Here are some things we pull: when you entered data, what you clicked on, when you clicked on it, and how long you had something open. We cannot yet read free text data like the contents of notes. Diagnosis groupings come from the CCSR standard.

### My Last 6 Months

#### Top Diagnoses Seen

| Diagnosis | Count |
|---|---|
| Respiratory Distress | 56 |
| Malnutrition / Fail to thrive | 48 |
| Bronchiolitis | 36 |
| Gastroenteritis | 33 |
| Skin/Soft Tissue Infection | 17 |
| Epilepsy / Seizure | 8 |
| Major Depressive D/O | 5 |
| Pneumonia | 5 |
| Asthma | 4 |
| Urinary Tract Infection | 4 |

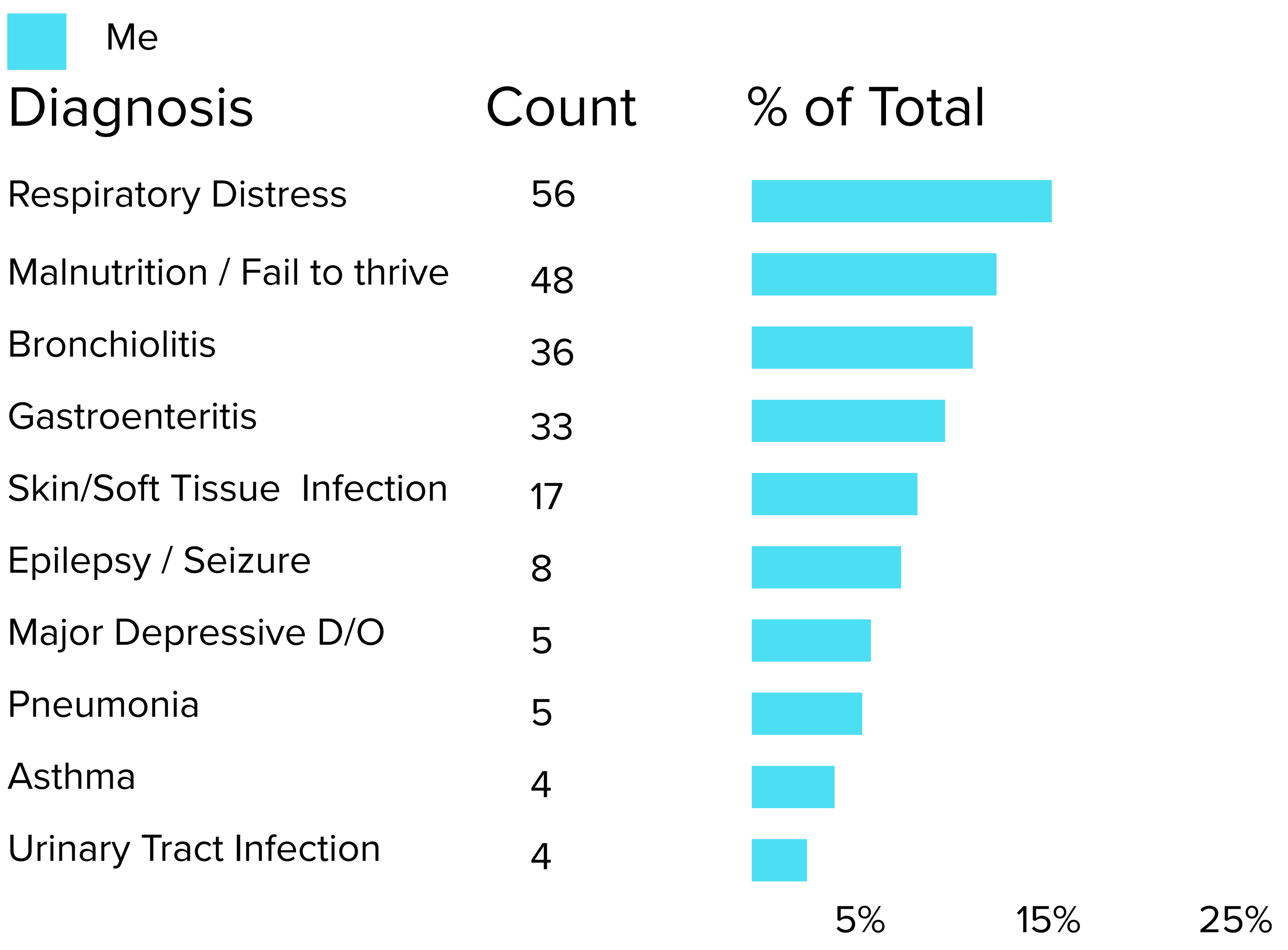


#### Gaps in Diagnosis Exposure

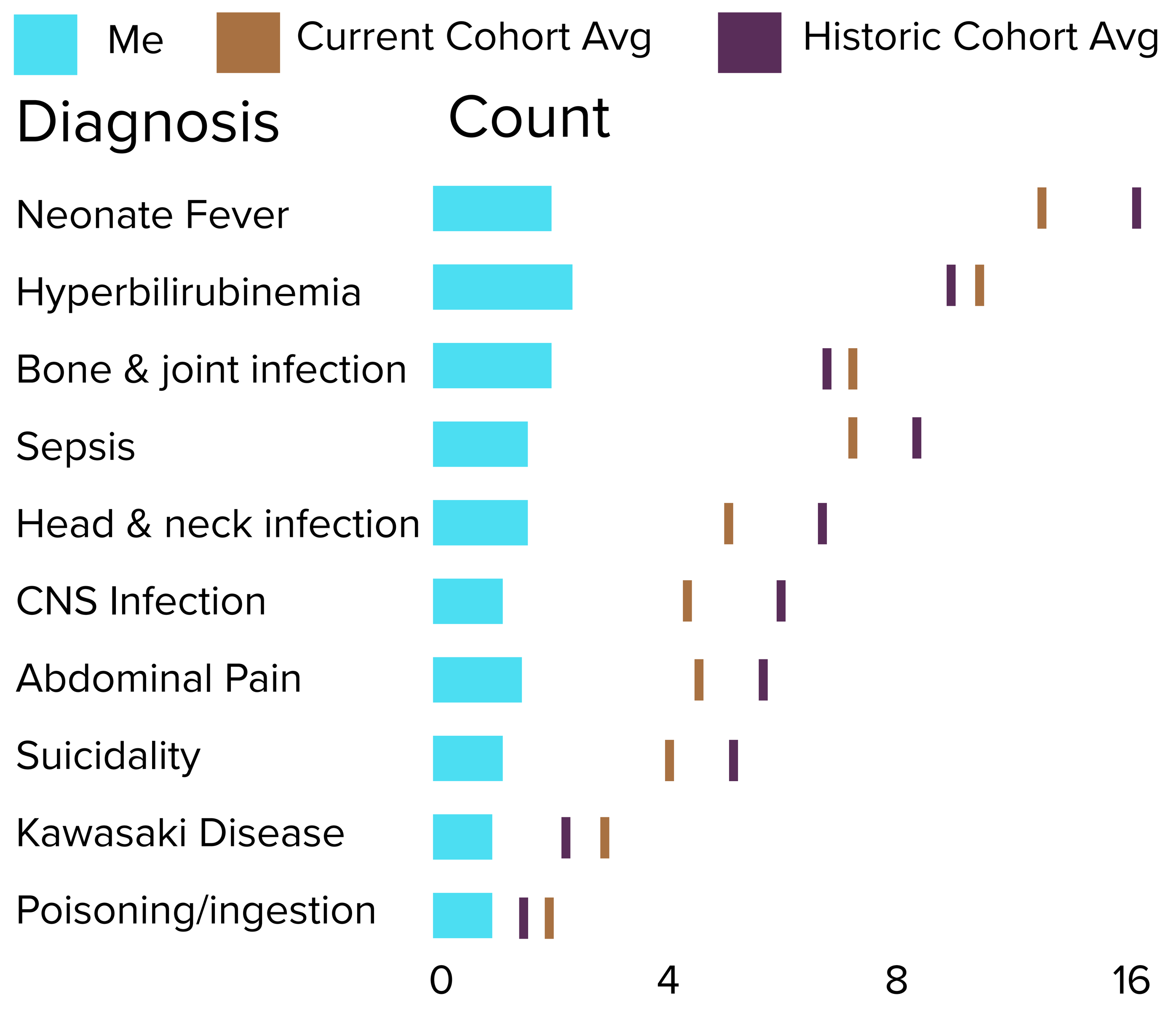


### What is my exposure to acuity and complex care ?

#### My Patient Makeup

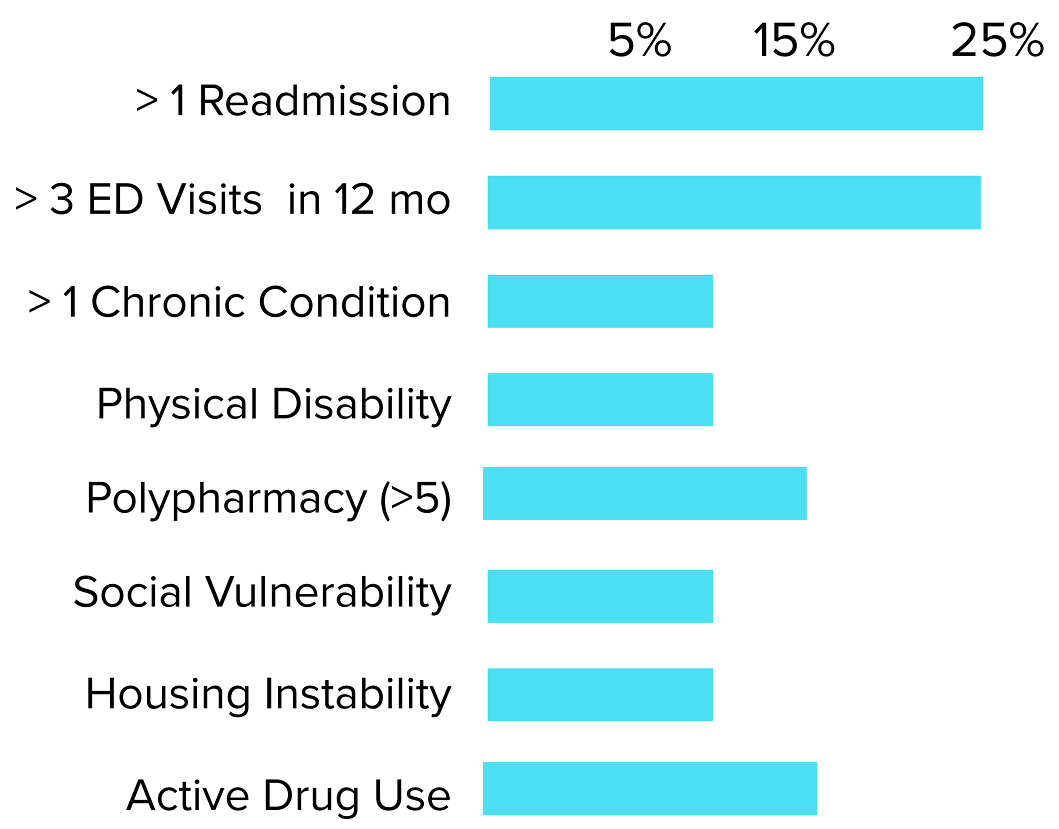

### Diagnosis Breakdown

#### What diagnoses am I seeing most?

Based on problem list data over the last 6 months

Non ED  Acuity Level 5  Acuity Level 4  Acuity Level 3  Acuity Level 2  Acuity Level 1

#### # 1 Respiratory Distresss

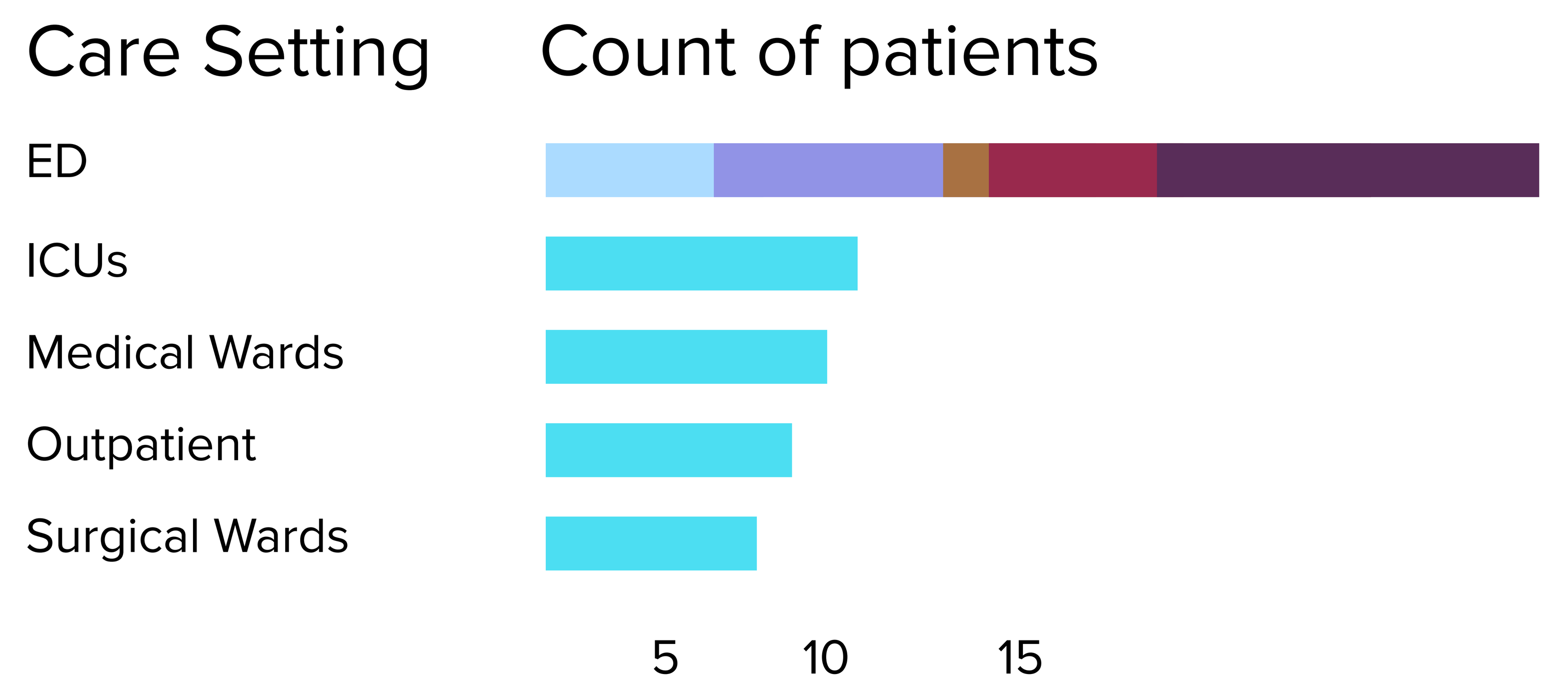


#### # 2 Malnutrition / Fail to Thrive

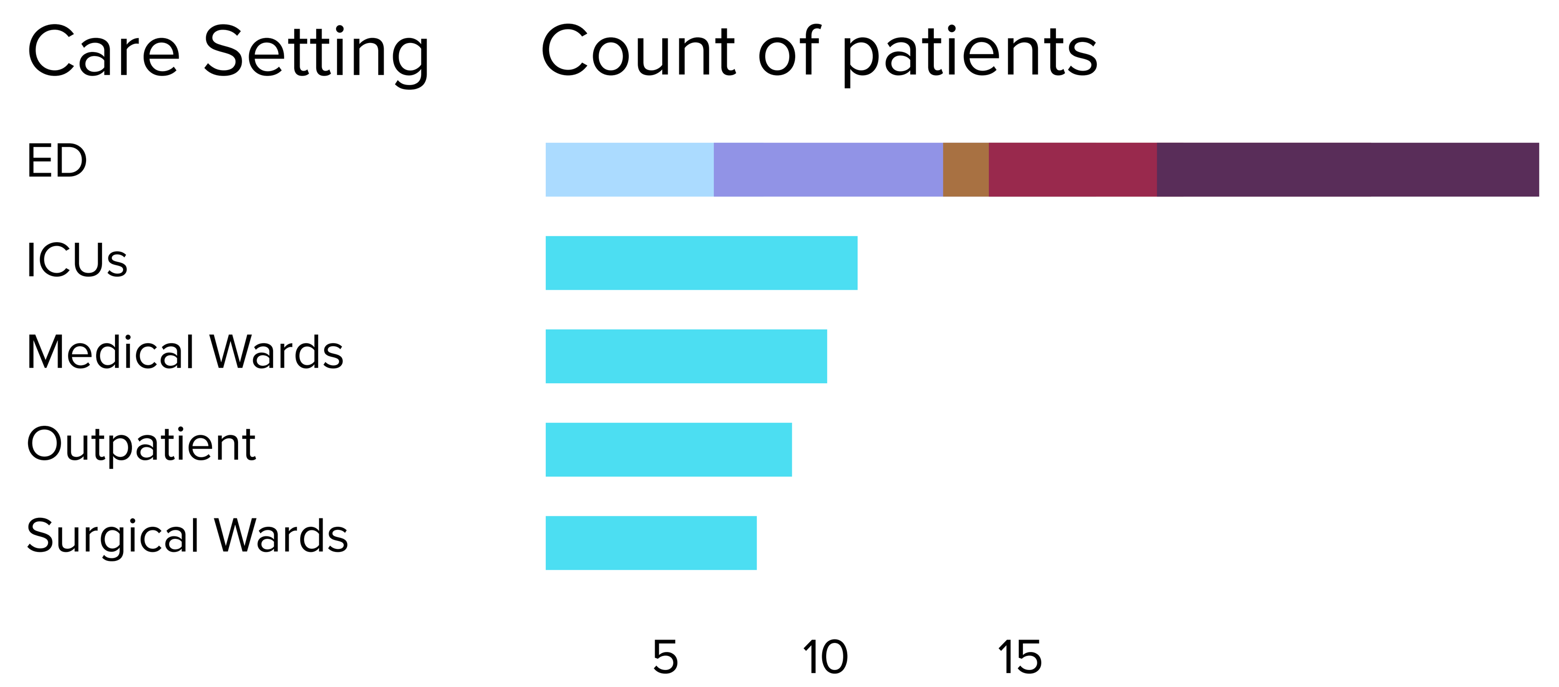


#### # 3 Bronchiolitis

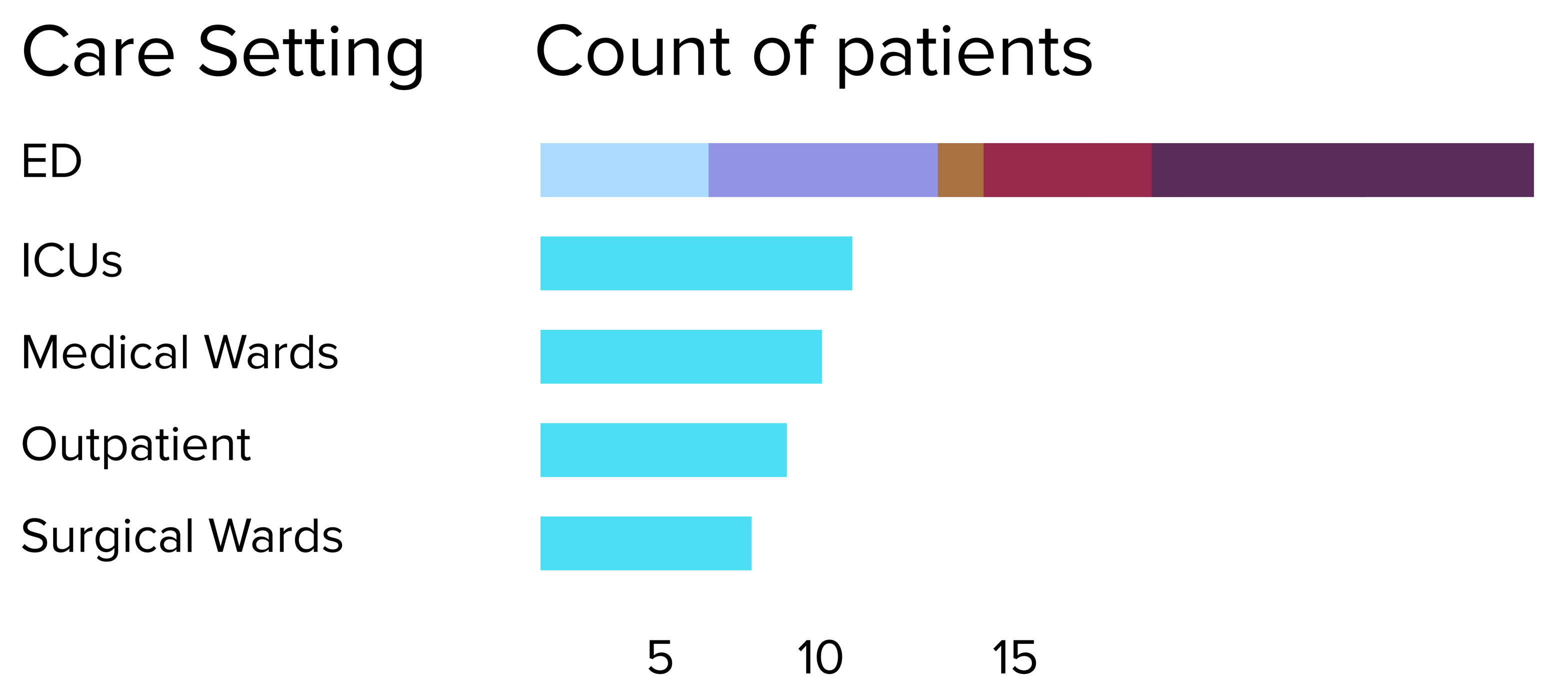


#### # 4 Gastroenteritis

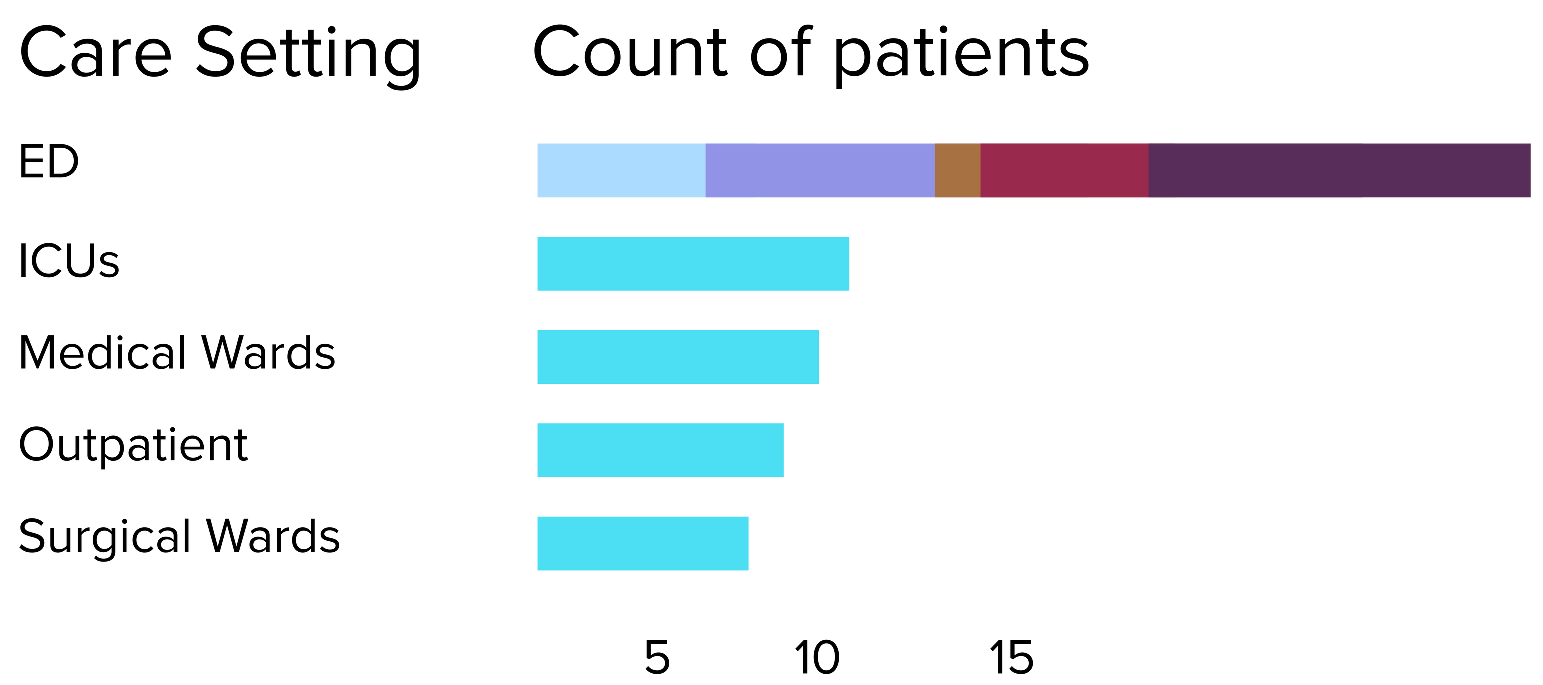


#### # 5 Skin and Soft Tissue Infection

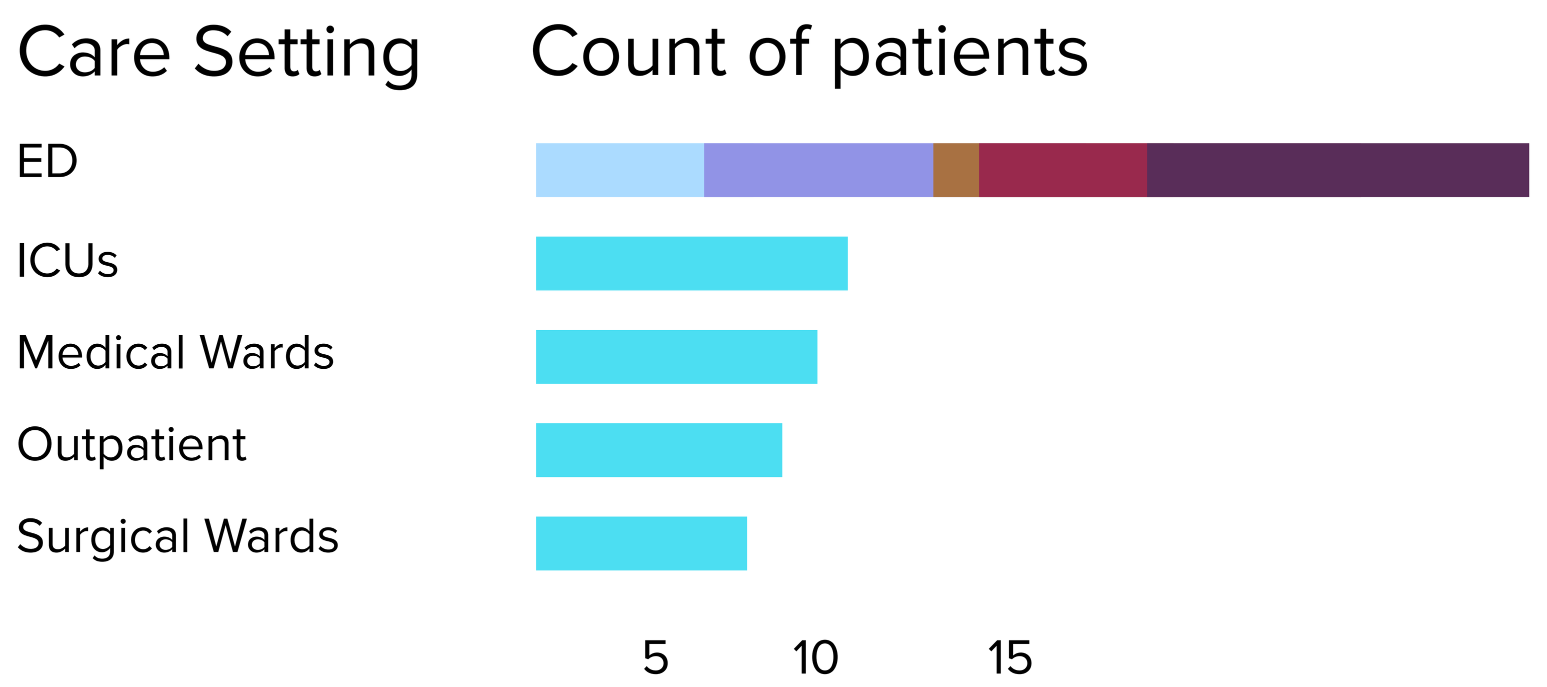


#### # 6 Epilepsy / Seizure

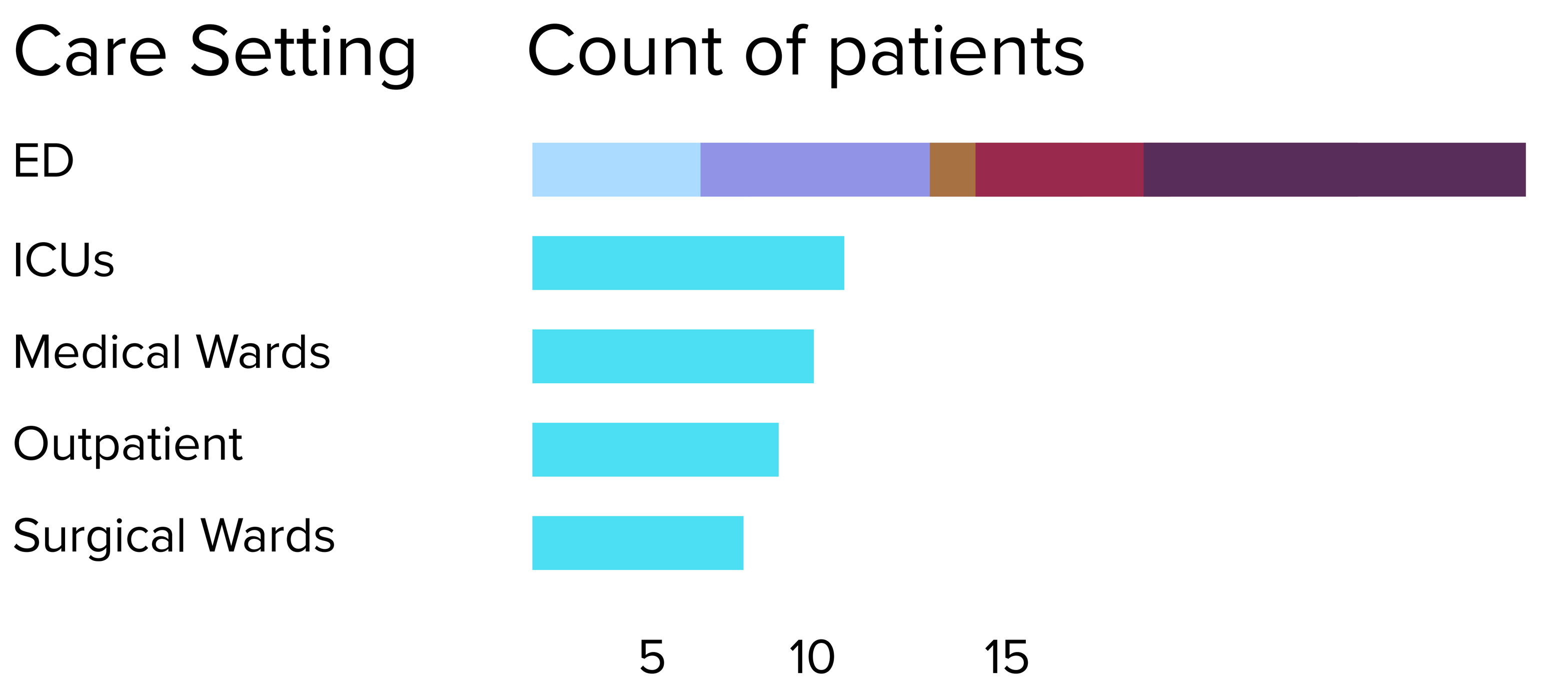


#### # 7 Mental Health

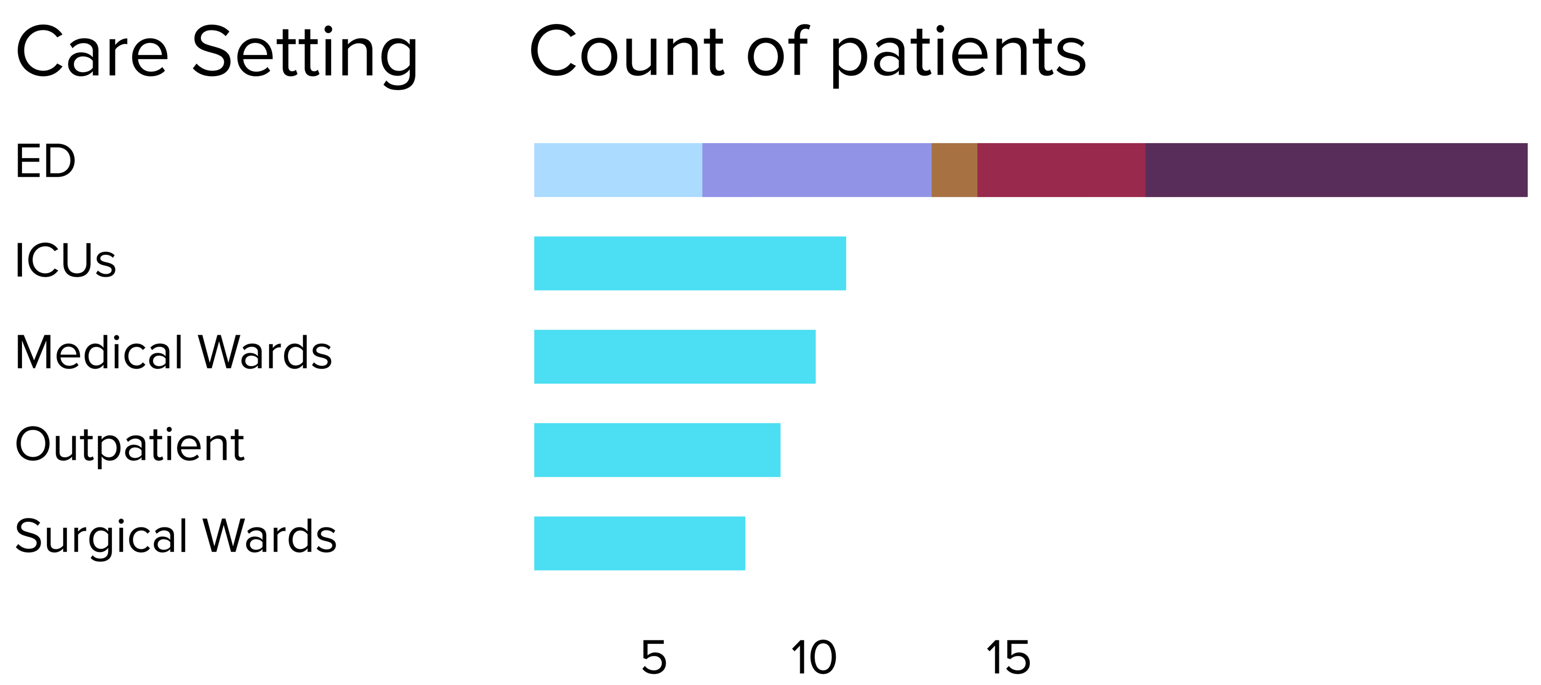


#### # 8 Pneumonia

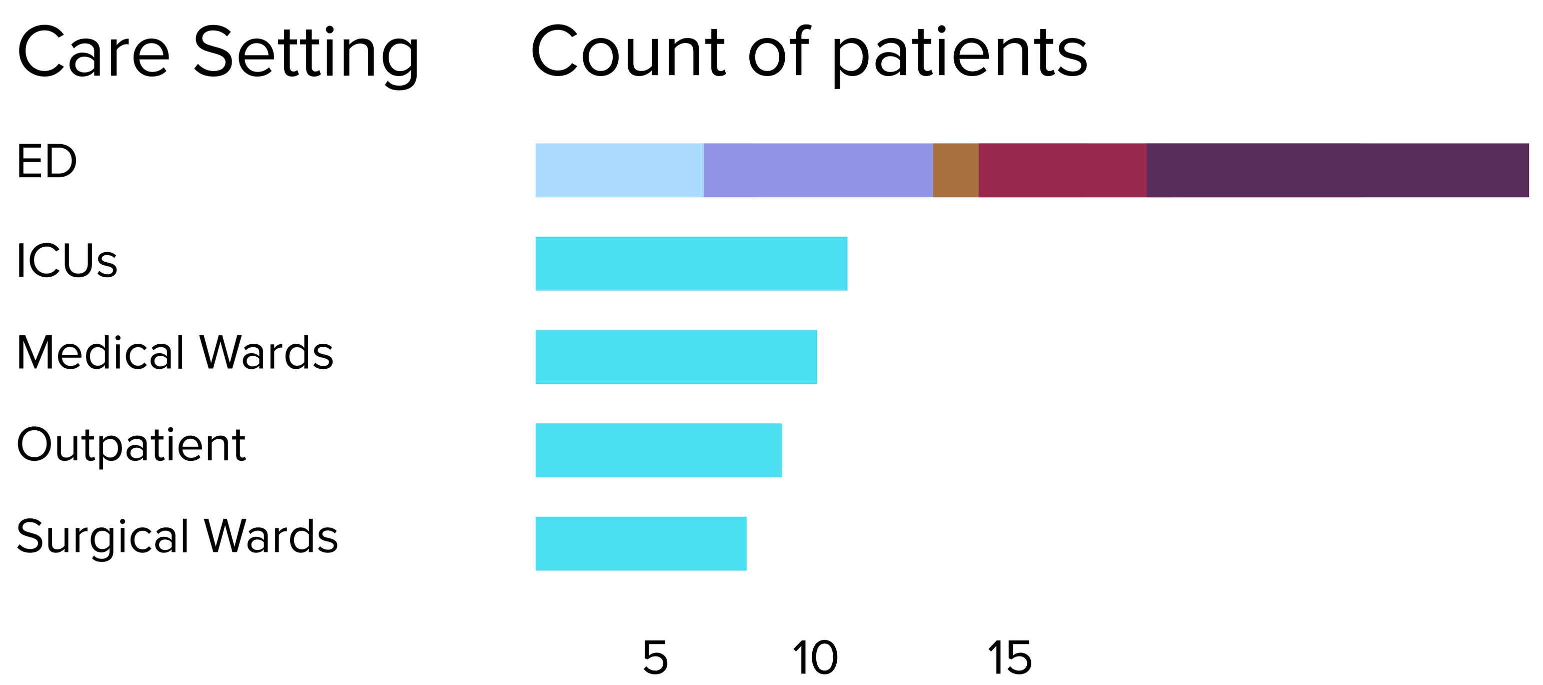

Electives to consider

### Pulmonary Medicine Elective

#### How do I read this part of the report?

Data shown in blue tells what your practice areas would look like if you took this particular elective.

This data comes from the last 6 months on this rotation. Where possible, it focuses on what other PGY-? experienced.

Compare the blue data to your own, in brown. Your data comes from activity in the last 6 months.

Compare the two and **decide if this elective matches your needs**, exposing you to new practice areas in settings that work with your schedule.

#### Will I see important diagnoses?

##### Top diagnoses by Δ of elective makeup vs my past patient makeup

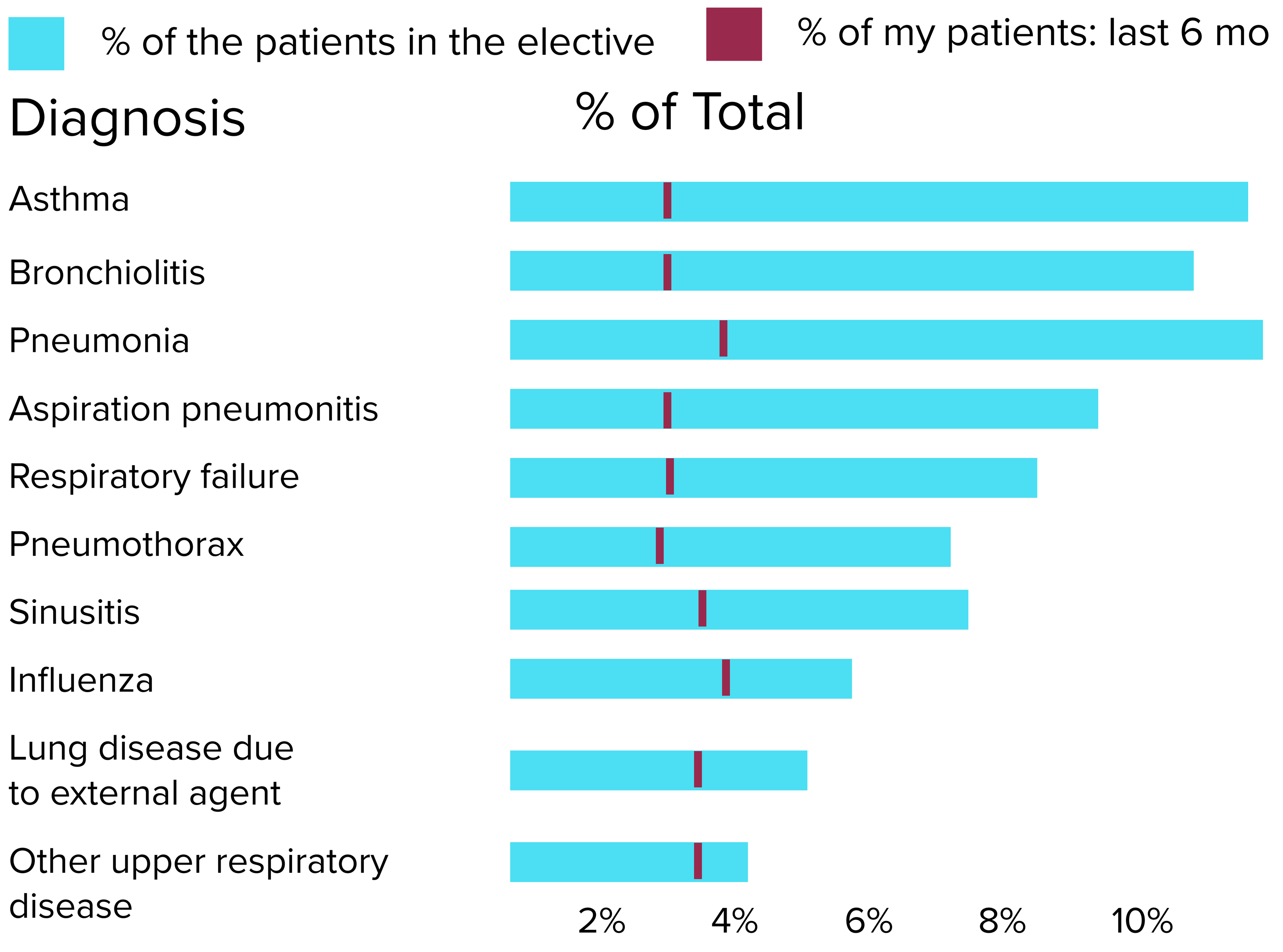

Electives to consider

### Neurology Elective

#### How do I read this part of the report?

Data shown in blue tells what your practice areas would look like if you took this particular elective.

This data comes from the last 6 months on this rotation. Where possible, it focuses on what other PGY-? experienced.

Compare the blue data to your own, in brown. Your data comes from activity in the last 6 months.

Compare the two and **decide if this elective matches your needs**, exposing you to new practice areas in settings that work with your schedule.

#### Will I see important diagnoses?

##### Top diagnoses by Δ of elective makeup vs my past patient makeup

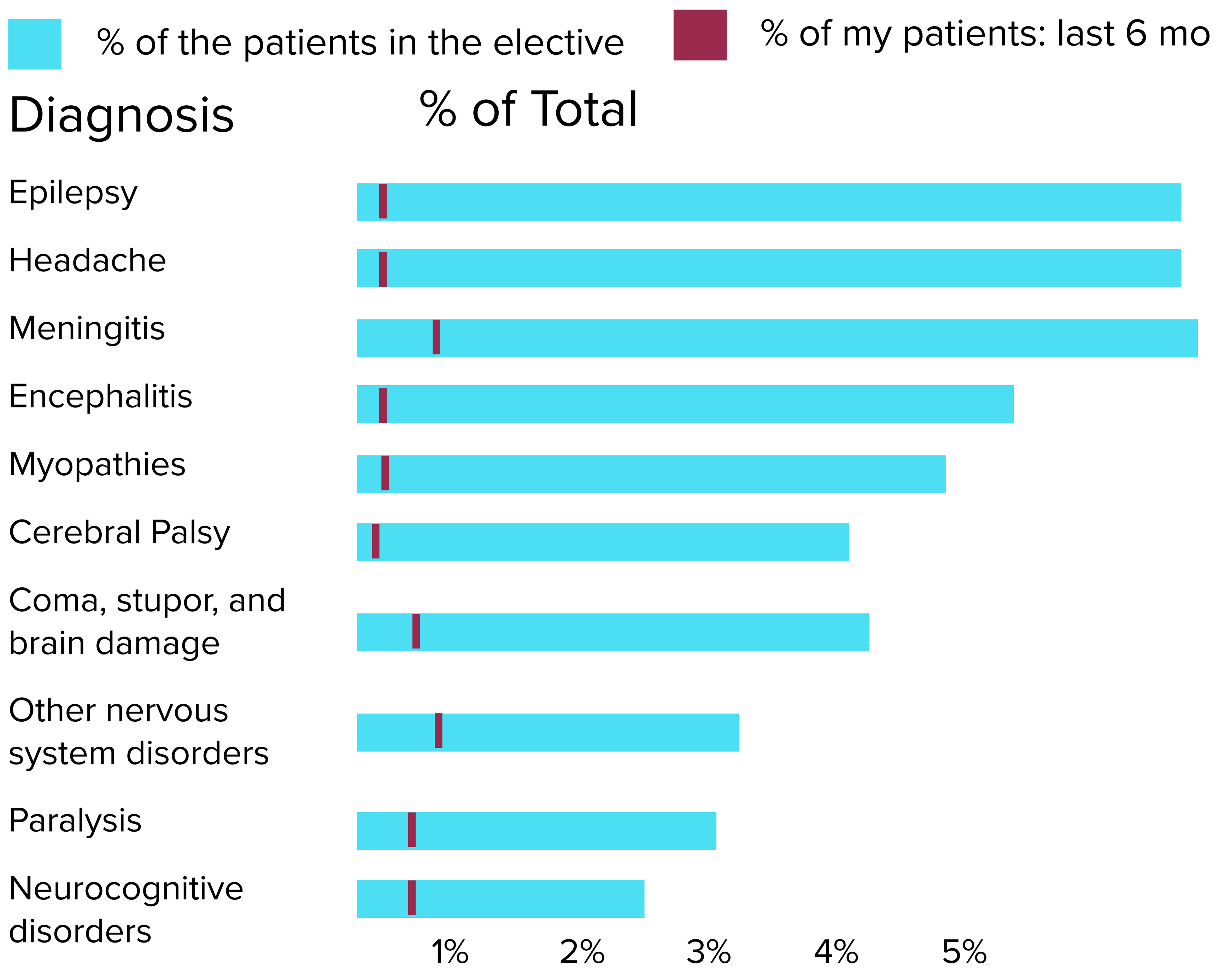

Electives to consider

### Infectious Disease Elective

#### How do I read this part of the report?

Data shown in blue tells what your practice areas would look like if you took this particular elective.

This data comes from the last 6 months on this rotation. Where possible, it focuses on what other PGY-? experienced.

Compare the blue data to your own, in brown. Your data comes from activity in the last 6 months.

Compare the two and **decide if this elective matches your needs**, exposing you to new practice areas in settings that work with your schedule.

#### Will I see important diagnoses?

##### Top diagnoses by Δ of elective makeup vs my past patient makeup

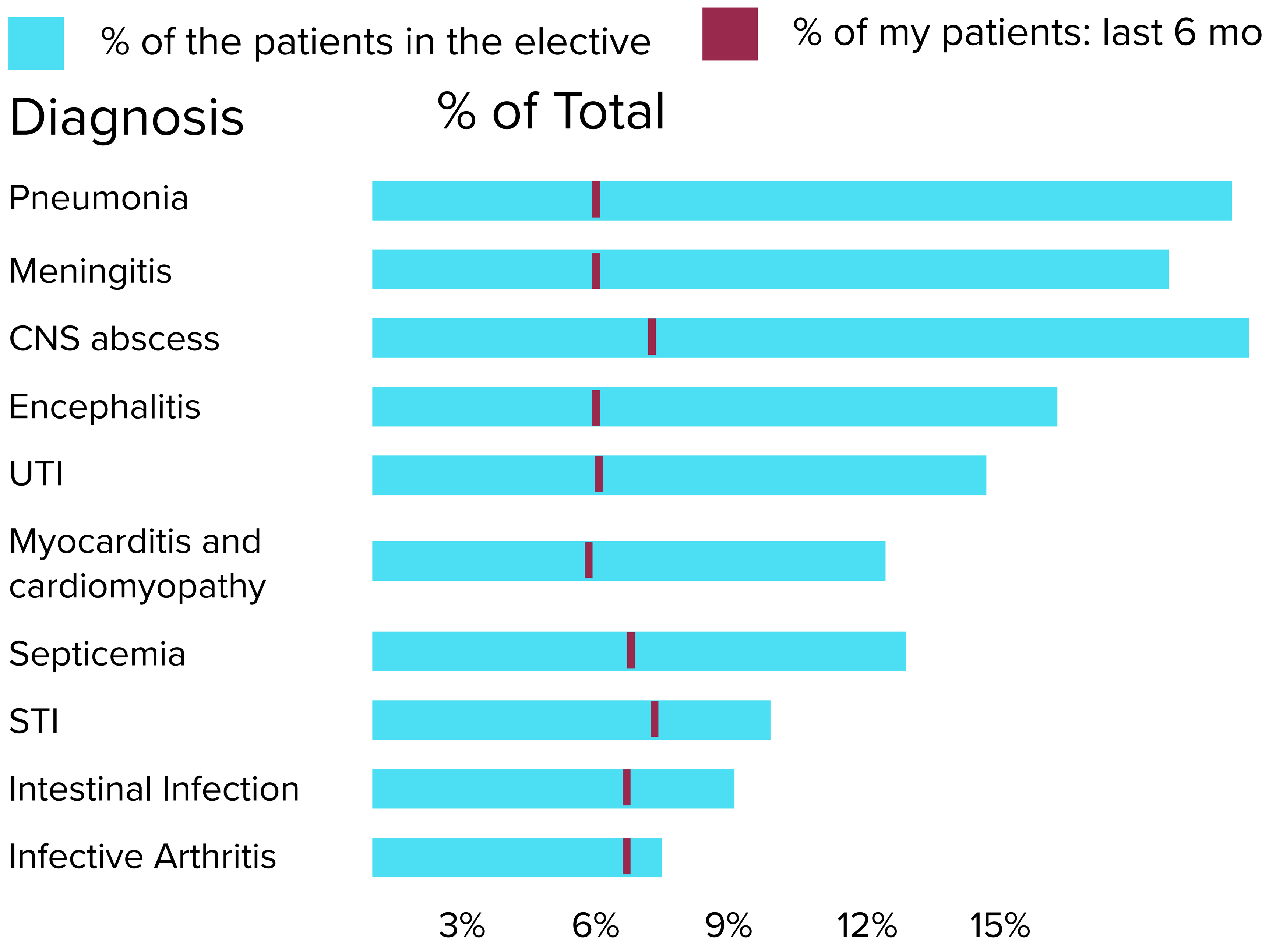
