## Supplementary material for "Precision Education Tools for Pediatrics Trainees: A Mixed-Methods Multi-Site Usability Assessment": REDCap UI Test

### Usability Review

Thank you for helping out with this important design research!

---

TRAILS Usability Test   Participant Instructions Download the report attached here if you do not alreay have a copy pulled up or printed out.

If viewing the report on your screen, please share that window or your whole screen.

Take 10 minutes to read the TRAILS report.

Use the TRAILS Report to answer questions. There are questions for 5 scenarios and a final review.

As you read the report and answer questions, think out loud, narrating your reactions and decisions.

The TRAILS Report TRAILS is a tool designed to support adaptive learning in trainees. It was designed to help you identify gaps between "what is" and "what could/should be" and, to then help you select a learning opportunity to close that gap.

[Attachment: "TRAILS Report for UI Test.pdf"]

---

1) Please check the boxes to move forward, showing that you...

- ☐ Read the report
- ☐ Still have the report open or printed
- ☐ Understand the "think aloud" protocol
- ☐ Are sharing the window or screen with the report, if you are reading it on screen.

#### Scenario 1 of 5

2) In the below scenario, when managing care of the three patients, for which patient could you ask for more autonomy?

Put another way, for which of the three patients could you consider moving up in entrustment milestones (seen below)?
Milestone 1: Observation only
Milestone 2: Execution with direct, proactive supervision
Milestone 3: Execution with direct, reactive supervision
Milestone 4: Supervision at a distance and/or post hoc
Milestone 5: Trainee supervises more junior colleagues

Scenario 1

You are on a general pediatrics inpatient service with three patients assigned to you. They have the following problem lists:

(Patient 1) RSV Bronchiolitis, Hydronephrosis, Feeding difficulties, Term birth of newborn female;

(Patient 2) Dehydration, Decreased oral intake; Hypernatremia;

(Patient 3) Premature infant of 29 weeks gestation; Failure to thrive; Sacral dimple in newborn; Abnormal movements; Delayed developmental milestones

- ◯ Patient 1
- ◯ Patient 2
- ◯ Patient 3

3) ______________________________

**Please rate the scenario and question you just finished.**

**1 is the strongest disagreement.**

**9 is the strongest agreement.**

**--**

**--**

| | | Strongly Disagree 1 | 2 | Moderately Disagree 3 | 4 | Undecided 5 | 6 | Moderately Agree 7 | 8 | Strongly Agree 9 |
|---|---|---|---|---|---|---|---|---|---|---|
| 4) | This scenario is important in finding learning or entrustment gaps and resolving them. | ○ | ○ | ○ | ○ | ○ | ○ | ○ | ○ | ○ |
| 5) | This scenario is realistic in the search for learning or entrustment gaps and resolving them. | ○ | ○ | ○ | ○ | ○ | ○ | ○ | ○ | ○ |
| 6) | In this scenario, the system was easy to use | ○ | ○ | ○ | ○ | ○ | ○ | ○ | ○ | ○ |
| 7) | In this scenario, the system allowed me to perform tasks efficiently | ○ | ○ | ○ | ○ | ○ | ○ | ○ | ○ | ○ |
| 8) | In this scenario, the system provided useful features | ○ | ○ | ○ | ○ | ○ | ○ | ○ | ○ | ○ |
| 9) | In this scenario, the system provided useful information | ○ | ○ | ○ | ○ | ○ | ○ | ○ | ○ | ○ |
| 10) | Overall, I am satisfied with how the system is designed in this scenario | ○ | ○ | ○ | ○ | ○ | ○ | ○ | ○ | ○ |
| 11) | In this scenario, the system is an improvement over what I would have used before | ○ | ○ | ○ | ○ | ○ | ○ | ○ | ○ | ○ |

12) ________________________________

#### Scenario 2 of 5

13) Scenario 2

You are part of a journal club or have an evidence-based practice presentation. It's your turn to choose the article or clinical question.

What disease could you focus on that would boost your own competency where it might be lowest, and at the same time leverage already deep experience of your peers?

__________________________________

14)

__________________________________

**Please rate the scenario and question you just finished.**

**1 is the strongest disagreement.**

**9 is the strongest agreement.**

**--**

**--**

| | | Strongly Disagree 1 | 2 | Moderately Disagree 3 | 4 | Undecided 5 | 6 | Moderately Agree 7 | 8 | Strongly Agree 9 |
|---|---|---|---|---|---|---|---|---|---|---|
| 15) | This scenario is important in finding learning or entrustment gaps and resolving them. | ○ | ○ | ○ | ○ | ○ | ○ | ○ | ○ | ○ |
| 16) | This scenario is realistic in the search for learning or entrustment gaps and resolving them. | ○ | ○ | ○ | ○ | ○ | ○ | ○ | ○ | ○ |
| 17) | In this scenario, the system was easy to use | ○ | ○ | ○ | ○ | ○ | ○ | ○ | ○ | ○ |
| 18) | In this scenario, the system allowed me to perform tasks efficiently | ○ | ○ | ○ | ○ | ○ | ○ | ○ | ○ | ○ |
| 19) | In this scenario, the system provided useful features | ○ | ○ | ○ | ○ | ○ | ○ | ○ | ○ | ○ |
| 20) | In this scenario, the system provided useful information | ○ | ○ | ○ | ○ | ○ | ○ | ○ | ○ | ○ |
| 21) | Overall, I am satisfied with how the system is designed in this scenario | ○ | ○ | ○ | ○ | ○ | ○ | ○ | ○ | ○ |
| 22) | In this scenario, the system is an improvement over what I would have used before | ○ | ○ | ○ | ○ | ○ | ○ | ○ | ○ | ○ |

23) ______________________________

#### Scenario 3 of 5

24) In the below scenario, for which patient would you want to seek more help or supervision?

- ◯ Patient 1
- ◯ Patient 2
- ◯ Patient 3

Scenario 3

You are on service in the PICU with three patients assigned to you where they have the following problem lists. Again, you are on service in the PICU.

(1) A 10yo F with severe malnutrition, chronic respiratory failure requiring a tracheostomy and ventilator, global developmental delay and seizures, with gastroenteritis and feeding intolerance;

(2) A 14yo M with obstructive sleep apnea and dysphagia, here with status asthmaticus;

(3) A 4yo F with a mitochondrial disease, abnormal movements and developmental delay, here with pneumonia and a complex pleural effusion.

25) ______________________________

**Please rate the scenario and question you just finished.**

**1 is the strongest disagreement.**
**9 is the strongest agreement.**
**--**
**--**

| | | Strongly Disagree 1 | 2 | Moderately Disagree 3 | 4 | Undecided 5 | 6 | Moderately Agree 7 | 8 | Strongly Agree 9 |
|---|---|---|---|---|---|---|---|---|---|---|
| 26) | This scenario is important in finding learning or entrustment gaps and resolving them. | ○ | ○ | ○ | ○ | ○ | ○ | ○ | ○ | ○ |
| 27) | This scenario is realistic in the search for learning or entrustment gaps and resolving them. | ○ | ○ | ○ | ○ | ○ | ○ | ○ | ○ | ○ |
| 28) | In this scenario, the system was easy to use | ○ | ○ | ○ | ○ | ○ | ○ | ○ | ○ | ○ |
| 29) | In this scenario, the system allowed me to perform tasks efficiently | ○ | ○ | ○ | ○ | ○ | ○ | ○ | ○ | ○ |
| 30) | In this scenario, the system provided useful features | ○ | ○ | ○ | ○ | ○ | ○ | ○ | ○ | ○ |
| 31) | In this scenario, the system provided useful information | ○ | ○ | ○ | ○ | ○ | ○ | ○ | ○ | ○ |
| 32) | Overall, I am satisfied with how the system is designed in this scenario | ○ | ○ | ○ | ○ | ○ | ○ | ○ | ○ | ○ |
| 33) | In this scenario, the system is an improvement over what I would have used before | ○ | ○ | ○ | ○ | ○ | ○ | ○ | ○ | ○ |

34) ______________________________

#### Scenario 4 of 5

35) Compared to the others, which patient could you could ask for more autonomy or entrustment in dealing with?

- ◯ Patient 1
- ◯ Patient 2
- ◯ Patient 3

Scenario 4

You are on service on the Neurology inpatient service with three patients assigned to you:

(Patient 1) A 13yo with debilitating migraine headaches

(Patient 2) A 7mo recovering from bacterial meningitis with auditory dysfunction

(Patient 3) A 4yo with developmental delay, refractory epilepsy, myoclonus, frequent urinary tract infections, failure to thrive, skin breakdown, presenting with worsening seizure frequency in the setting of a respiratory viral illness.

---

36) ________________________________

**Please rate the scenario and question you just finished.**

**1 is the strongest disagreement.**

**9 is the strongest agreement.**

**--**

**--**

| | | Strongly Disagree 1 | 2 | Moderately Disagree 3 | 4 | Undecided 5 | 6 | Moderately Agree 7 | 8 | Strongly Agree 9 |
|---|---|---|---|---|---|---|---|---|---|---|
| 37) | This scenario is important in finding learning or entrustment gaps and resolving them. | ○ | ○ | ○ | ○ | ○ | ○ | ○ | ○ | ○ |
| 38) | This scenario is realistic in the search for learning or entrustment gaps and resolving them. | ○ | ○ | ○ | ○ | ○ | ○ | ○ | ○ | ○ |
| 39) | In this scenario, the system was easy to use | ○ | ○ | ○ | ○ | ○ | ○ | ○ | ○ | ○ |
| 40) | In this scenario, the system allowed me to perform tasks efficiently | ○ | ○ | ○ | ○ | ○ | ○ | ○ | ○ | ○ |
| 41) | In this scenario, the system provided useful features | ○ | ○ | ○ | ○ | ○ | ○ | ○ | ○ | ○ |
| 42) | In this scenario, the system provided useful information | ○ | ○ | ○ | ○ | ○ | ○ | ○ | ○ | ○ |
| 43) | Overall, I am satisfied with how the system is designed in this scenario | ○ | ○ | ○ | ○ | ○ | ○ | ○ | ○ | ○ |
| 44) | In this scenario, the system is an improvement over what I would have used before | ○ | ○ | ○ | ○ | ○ | ○ | ○ | ○ | ○ |

45) ______________________________

#### Scenario 5 of 5

46) Scenario 5

You must chose one of the three electives.

Which will expose you to diagnosis and management of diseases least familiar to you?

- ◯ Pulmonary Medicine
- ◯ Neurology
- ◯ Infectious Disease

47) ________________________________

**Please rate the scenario and question you just finished.**

**1 is the strongest disagreement.**

**9 is the strongest agreement.**

**--**

**--**

| | | Strongly Disagree 1 | 2 | Moderately Disagree 3 | 4 | Undecided 5 | 6 | Moderately Agree 7 | 8 | Strongly Agree 9 |
|---|---|---|---|---|---|---|---|---|---|---|
| 48) | This scenario is important in finding learning or entrustment gaps and resolving them. | ◯ | ◯ | ◯ | ◯ | ◯ | ◯ | ◯ | ◯ | ◯ |
| 49) | This scenario is realistic in the search for learning or entrustment gaps and resolving them. | ◯ | ◯ | ◯ | ◯ | ◯ | ◯ | ◯ | ◯ | ◯ |
| 50) | In this scenario, the system was easy to use | ◯ | ◯ | ◯ | ◯ | ◯ | ◯ | ◯ | ◯ | ◯ |
| 51) | In this scenario, the system allowed me to perform tasks efficiently | ◯ | ◯ | ◯ | ◯ | ◯ | ◯ | ◯ | ◯ | ◯ |
| 52) | In this scenario, the system provided useful features | ◯ | ◯ | ◯ | ◯ | ◯ | ◯ | ◯ | ◯ | ◯ |
| 53) | In this scenario, the system provided useful information | ◯ | ◯ | ◯ | ◯ | ◯ | ◯ | ◯ | ◯ | ◯ |
| 54) | Overall, I am satisfied with how the system is designed in this scenario | ◯ | ◯ | ◯ | ◯ | ◯ | ◯ | ◯ | ◯ | ◯ |
| 55) | In this scenario, the system is an improvement over what I would have used before | ◯ | ◯ | ◯ | ◯ | ◯ | ◯ | ◯ | ◯ | ◯ |

56) ______________________________

#### Final Review of Overall System

Almost done! Last set of questions is on the next page. Thanks again for participating.

**For our last set of questions, please think about the report *as a whole*.**

**1 is the strongest disagreement.**
**9 is the strongest agreement.**
**--**
**--**

| | Strongly Disagree 1 | 2 | Moderately Disagree 3 | 4 | Undecided 5 | 6 | Moderately Agree 7 | 8 | Strongly Agree 9 |
|---|---|---|---|---|---|---|---|---|---|
| 57) Overall, the report was easy to use | ○ | ○ | ○ | ○ | ○ | ○ | ○ | ○ | ○ |
| 58) Overall, the report allowed me to perform tasks efficiently | ○ | ○ | ○ | ○ | ○ | ○ | ○ | ○ | ○ |
| 59) Overall, the report provided useful features | ○ | ○ | ○ | ○ | ○ | ○ | ○ | ○ | ○ |
| 60) Overall, the report provided useful patient information | ○ | ○ | ○ | ○ | ○ | ○ | ○ | ○ | ○ |
| 61) Overall, I am satisfied with how the report is designed to address the search for and resolution of gaps in learning and entrustment | ○ | ○ | ○ | ○ | ○ | ○ | ○ | ○ | ○ |
| 62) Overall, the report presents an improvement over existing tools to find and resolve gaps in learning and entrustment | ○ | ○ | ○ | ○ | ○ | ○ | ○ | ○ | ○ |
